# Low-grade inflammation and daily life food-related motivation in obesity

**DOI:** 10.64898/2026.06.26.26356659

**Authors:** Judith M. Scholing, Ruben van den Bosch, Lisette Olsthoorn, Julia C.J. Loenen, Catharina M. Mulders-Manders, Rinke Stienstra, Esther Aarts

**Affiliations:** Donders Institute for Brain, Cognition and Behaviour, Radboud University, Nijmegen, The Netherlands; Department of Internal Medicine, Radboud University Medical Center, Nijmegen, the Netherlands; Division of Human Nutrition and Health, Wageningen University C Research, Wageningen, the Netherlands

## Abstract

Obesity is associated with low-grade inflammation, which – our prior work shows – causally and reversibly increases effort aversion and related brain responses during food-related decision-making. However, how these laboratory findings translate to daily-life (food) motivation remains unclear. This study investigated low-grade inflammation in more ecologically valid measures of (food-related) motivation in obesity.

We conducted a cross-sectional study (N=145 women, BMI>27 kg/m2, 18-59 yrs) and a 12-week randomized controlled trial (N=57 women, BMI>30 kg/m2, 18-59 years, C-reactive protein (CPR)>3 mg/l) testing the anti-inflammatory drug colchicine versus placebo. We measured effort-related food intake using a bogus taste test, daily-life motivation using ecological momentary assessment (EMA), and dietary intake using a food frequency questionnaire (FFQ).

INFLA-score (CRP, white blood cell count, neutrophil-to-lymphocyte ratio, platelets) related to lower intake of high-effort food items on the taste test (β –0.26 SD, p=0.011), lower engagement in high-effort activities (OR 0.72, p<0.001), and negatively moderated the association between anticipation and activity completion in EMA (OR 0.89, p=0.032). On the FFQ, INFLA-score related to lower intake of legumes and fruits and more intake of processed meat (all p<0.05). Colchicine decreased overall caloric intake (β –341 kcal, p=0.018). A decrease in inflammation related to healthier dietary choices (Spearman ρ=0.37, p=0.013). Colchicine did not affect effort-related food intake on the taste test or EMA.

These findings indicate that that obesity-related inflammation is associated with increased effort-related food intake and motivation in daily life, and that reducing inflammation decreases (unhealthy) food intake. Such effects may partly underlie the challenges of achieving and maintaining weight loss.

## Introduction

The prevalence of obesity is increasing worldwide, yet the mechanisms contributing to its development and persistence are not well understood. One such mechanism may be chronically increased inflammation levels often present in individuals with obesity. This state of metabolic inflammation can dysregulate key hormonal processes involved in energy balance, thereby making weight loss more difficult (AL-Suhaimi and Shehzad 2013; Tchernof and Després 2013). In addition, we demonstrated that this systemic, low-grade inflammation in obesity alters food choice and (health-related) motivation. Specifically, low-grade inflammation lowered motivated behaviour by causing effort aversion (Scholing et al., preprint). This could further contribute to difficulties in initiating and maintaining lifestyle changes and weight loss, as reflected in lower participation and completion rates of lifestyle programs in individuals with higher BMI shown in previous studies (Burgess, Hassmén, and Pumpa 2017; Greenberg et al. 2009; Lemstra et al. 2016).

In the context of obesity, one particularly relevant type of behaviour driven by motivational processes is effort-based decision making. For example, food choices can be conceptualised as a form of effort-based decision-making, where individuals assess whether the type of food is worth expending the required effort for (Chong, Bonnelle, and Husain 2016). These decisions are driven by motivational processes, particularly one’s aversion to effort and one’s sensitivity to rewards. An imbalance in these two processes can lead to maladaptive decision-making, such as preference for easily-available high caloric foods (Romieu et al. 2017). Obesity has been associated with altered willingness to exert effort for rewards compared to individuals with normal weight, indicating such an imbalance between effort and reward sensitivity (Epstein et al. 2007; Giesen et al. 2010; Mata et al. 2017; Mathar et al. 2016; Miras et al. 2012; Rollins et al. 2014). Identifying the factors that contribute to these motivational imbalances will help provide insight into the behavioural mechanisms that sustain obesity.

Systemic, low-grade inflammation may explain these differences in motivated behaviour. Obesity is characterized by low-grade inflammation, driven by macrophage infiltration into adipose tissue and pro-inflammatory dietary patterns (Buyken et al. 2014; Rohm et al. 2022; Saltiel and Olefsky 2017; Shivappa et al. 2014; Zhou, Urso, and Jadeja 2020). Outside the research field of obesity, studies show that acute inflammation typically induces sickness behaviour, characterized by low motivation, anhedonia and fatigue (Dantzer 2001; De Marco et al. 2023; Lasselin et al. 2017). More specifically, inducing acute inflammation in healthy volunteers with lipopolysaccharide increased effort aversion during effort-based decision-making tasks, without affecting reward sensitivity (Draper et al. 2018; Lambregts et al. 2023; Lasselin et al. 2017). In line with these acute inflammatory effects, our recent work reveals that low-grade inflammation also increases effort aversion during an effort-based decision-making task in obesity (Scholing, et al., 2026 preprint). This indicates that obesity-related systemic, low-grade inflammation may be an important factor in the maladaptive motivated behaviour observed in obesity.

To date, our work and much of the existing literature on the effects of inflammation on decision-making has relied on computerized behavioural tasks conducted in laboratory settings, which offer limited ecological validity. In the context of obesity, Mata and colleagues showed that less willingness to expend effort for rewards during a computerized behavioural task related to poorer adherence to a weight loss intervention, indicating that reduced effort-related motivation in laboratory tasks may translate into lower motivation in daily life (Mata et al. 2017). Despite this, evidence on the role of inflammation in motivated behaviour in daily life settings is currently lacking. Consequently, it remains unclear how the inflammation-related alterations in motivation translate to motivated behaviour outside the laboratory.

The objective of this study was to investigate the role of low-grade inflammation on motivated behaviour in daily life of women with a BMI>27 kg/m^2^. Based on our recent findings of inflammation increasing effort aversion in a laboratory setting (Scholing et al. 2026 preprint), we hypothesized that obesity-related inflammation leads to less choice of high-effort food items and less effort exertion in daily life.

We conducted a cross-sectional study to investigate the association between circulating inflammatory markers and motivation in daily life. In addition, we performed a randomized controlled trial to study the effects of lowering inflammation using colchicine versus placebo. To study motivated behaviour, we used more ecologically valid measures than a computerized task: a newly-designed bogus food taste test to assess effort-related food intake, an ecological momentary assessment (EMA) to assess active and effortful behaviour at home, and a food frequency questionnaire to assess general diet quality in the last month.

## Methods

### Study design G population

Participants were recruited from social media and local newspaper advertisements, as well as from the Radboud University’s participant databases. Participants were eligible if they were aged between 18-59 years, had a BMI >27 kg/m2, had not been sick or vaccinated in the 4 weeks prior to participation, did not have inflammation levels indicating acute inflammation (CRP>10 mg/L for BMI ≤31 kg/m^2^ and CRP>22.1 mg/L for BMI >31 kg/m^2^), and did not have diabetes, autoimmune, psychiatric, or inflammatory disease.

Participants attended a study visit, during which they performed a bogus food taste test, MRI measurement and behavioural tests (Fig. 1; Scholing et al, preprint). In addition, a blood sample was taken. In the week following the study visit, participants completed an EMA protocol of 10 days and a food frequency questionnaire at home.

**Fig. 1.**
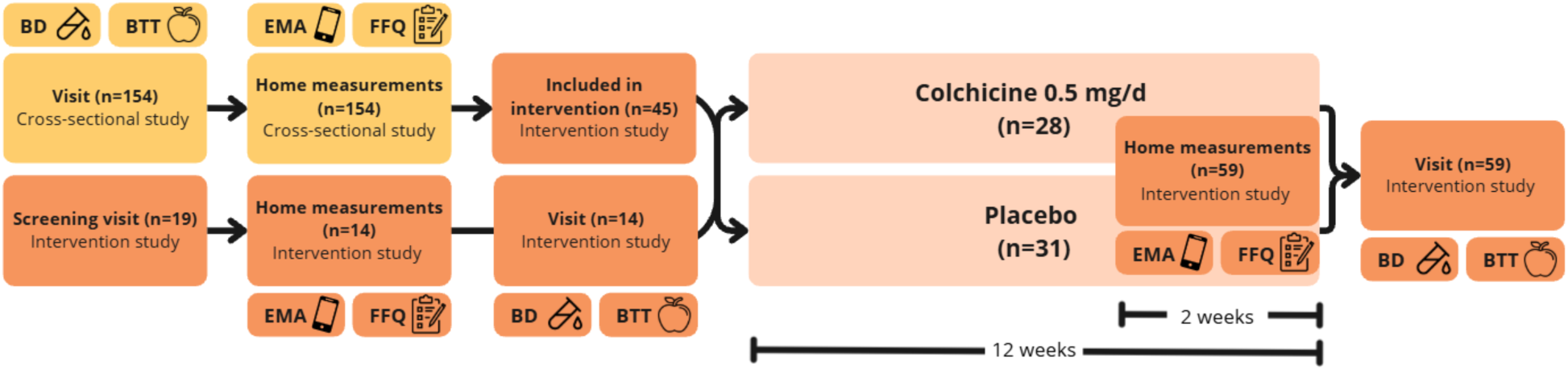
Study design. The cross-sectional study is illustrated in yellow, and the intervention study in orange. BD, blood drawing; BTT, bogus taste test; EMA, ecological momentary assessment; FFQ, food frequency questionnaire.

Participants with low-grade inflammation (CRP >3 mg/L) and a BMI >30 kg/m^2^ were invited to participate in a double-blind placebo-controlled randomized controlled trial (pre-registration: https://osf.io/2c3ax; Eudra-CT identifier: 2021-004919-11; ClinicalTrials.gov identifier: NCT05785429), during which they received either one tablet of 0.5 mg colchicine or a placebo for 12 weeks. Participants were randomized using a computer-generated randomization sequence (Castor Electronic Data Capture, Castor EDC), stratified by baseline age (18–40 vs. 41–59 years), BMI (30–35 vs. >35 kg/m²), and CRP levels (3.0–6.5 vs. >6.5 mg/L). In the final week of the intervention period, participants completed a second EMA protocol of 10 days and FFQ. After the study period was completed, participants attended a follow-up visit identical to the baseline study visit.

Both studies were approved by the Medical Ethics Review Committee Oost-Nederland, Nijmegen, the Netherlands (cross-sectional study: NL77503.091.21; intervention study: NL79408.091.21). All study procedures were conducted in accordance with the Declaration of Helsinki and written informed consent was obtained from all participants prior to participation.

### Inflammation

We used the INFLA-score as an index of low-grade inflammation, which is a composite score including C-reactive protein, white blood cell count, neutrophil/lymphocyte ratio and platelets. The specific methods of the collection and analyses of these inflammatory markers are described elsewhere (Scholing et al., preprint; Bonaccio et al., 2016). The INFLA-score was computed by Z-scoring each of the four inflammatory markers and averaging these standardized values.

### Bogus taste test

Effort– and reward-related food intake was measured using a bogus taste test (Robinson et al. 2017). Participants were presented with eight food items in identical bowls arranged in a circle to avoid implying any order. Food items differed in effort (more vs. less chewing/peeling). To account for caloric content and preference differences, for both low-and high-effort products we included two high– and two low-caloric options and two sweet and two savoury options (Table 1). Participants were told that we were interested in how they experienced these different foods and were asked to taste and rate each food on liking, wanting, sweetness, saltiness, sourness, fattiness, texture and aftertaste on a 100-point Likert-scale. Participants were left alone for 20 minutes to complete the questionnaire, and were told that they could take as many bites as they wanted to complete the questions for each food item. Unbeknownst to the participants, we measured their food intake by measuring the weight of the bowls before and after the test. Participants with intolerances for certain products received a similar product in order to have 8 products again. However, the intake of these alternative products were excluded from the analysis due to caloric differences of these products.

**Table 1:** Food items included in the bogus taste test.

|  |  | Low effort | High effort |
| --- | --- | --- | --- |
| Low caloric | Sweet | Grapes | Apple |
|  | Savory | Cherry tomatoes | Carrots |
| High caloric | Sweet | Cake | Liquorice |
|  | Savory | Gouda cheese | Pistachios |

### Ecological momentary assessment

Motivated behaviour in daily life was measured using an EMA, based on a paradigm used to measure motivational dynamics in depressed patients (Bakker et al. 2017). During a maximum of 10 consecutive days, participants received 7 notifications per day to fill out a questionnaire via an app on their mobile phone (MovisensXS, movisens GmbH, Karlsruhe, Germany). After rating their mood, participants were asked to indicate what type of activity they were planning to do in the next hour (i.e. “eating or drinking – self prepared”, “eating or drinking – takeaway or eating out”, “eating or drinking – other”, “exercising or moving”, “working or studying”, “relaxing”, “doing groceries or shopping”, or “household tasks”), and how much they were looking forward to it on a 7-point Likert-scale. Finally, participants reported whether they had done what they had planned in the previous questionnaire, and to what extent they thought the activity was effortful (both 7-point Likert-scales). Participants continued the EMA measurement until they had filled out at least 49 questionnaires, or when they completed the measurement period of 10 days.

### Diet quality

To measure diet quality in daily life, we used the Dutch Healthy Diet index 2015 (DHD-index) as calculated from a 81-item Food-Frequency Questionnaire (FFQ) retrospectively measuring food intake in the last 30 days (Carver and White 1994; Eussen et al. 2018). The DHD-index indicates the adherence to the Dutch Dietary Guidelines by the Health Council of the Netherlands (0-160 pt), which is the sum of 16 components (fruit, vegetables) (Looman et al. 2017; de Rijk et al. 2021). We additionally included the extra unhealthy choices component, which includes foods high in energy, saturated fat and sugar that were not included in the other components (de Rijk et al. 2021). Furthermore, we used the FFQ to measure daily caloric, macronutrient and micronutrient intake. In this paper, we focus on the DHD-index, its subscores, and macronutrient intake as these are most relevant for measuring food choice behaviour in daily life.

### Mood, reward sensitivity, and general health questionnaires

We measured current mood and mood in the past month using the Profile of mood states (McNair et al. 1971) and the Hospital Anxiety and Depression Scale (Zigmond and Snaith 1983), respectively. We measured fatigue using the Checklist Individual Strength (The assessment of fatigue: Psychometric qualities and norms for the Checklist individual strength – PubMed n.d.), and apathy using the Starkstein Apathy scale (Starkstein et al. 1992). We studied motivation and reward sensitivity in daily life using the Behavioural Inhibition System/ Behavioural Activation System (BIS/BAS) questionnaire (Carver and White 1994), the Temporary Experienced Pleasure Scale (Simon et al. 2018), and the Kirby Questionnaire (Kirby and Maraković 1996). We measure food reward and binge-eating behaviour using the Power of Food Scale (Lowe et al. 2009) and the Binge Eating Scale (The assessment of binge eating severity among obese persons – PubMed n.d.), respectively. We measured general health using the SF-36 (Ware and Sherbourne 1992) and sleep quality using the Pittsburgh Sleep index (Buysse et al. 1989).

## Statistical methods

### Data quality checks

We included participants in the analysis if they had data on any of the main outcome measures (i.e. bogus taste test, EMA and FFQ) in this study. We screened all continuous variables for outliers using a cut-off of >3 SD alongside visual inspection of histograms and boxplots. Values were excluded if they indicated measurement error or were considered biologically implausible.

All analyses were performed in R studio (version 2025.05.1). All mixed-effects regression models were estimated using the afex package (version 1.4.1; Singmann et al., 2025). All continuous variables included in regression models were standardized prior to analysis.

### Demographics

We summarized baseline characteristics for both the cross-sectional and intervention populations using mean+SD for normally distributed continuous variables, median (25^th^, 75^th^ percentile) for skewed continuous variables, and n (%) for categorical variables. We tested differences between the intervention groups using independent t-tests for normally distributed continuous variables, Mann-Whitney U-tests for skewed continuous variables, and chi-square tests for categorical variables.

### Inflammation

We assessed the effect of the intervention using a linear mixed regression model with INFLA-score as the outcome, Timepoint (baseline, follow-up), Group (colchicine, placebo) and their interaction as fixed factors, and Age and baseline BMI as covariates. A random intercept of participant and a random slope for Timepoint were included to account for within-subject variability.

### Inflammation and effort-related food intake in the bogus taste test

We investigated the relationship between INFLA-score and effort-related food intake by using a linear mixed linear regression model with caloric intake as outcome variable. The model included INFLA-score, effort (high, low), liking of the product and their interaction as fixed factors of interest.

The effect of colchicine on effort-related food intake was assessed using a linear mixed regression model with caloric intake as outcome variable, and Group (colchicine, placebo), Timepoint (baseline, follow-up), Effort (high, low) and Liking and their interaction as fixed factors of interest. Furthermore, we studied whether changes in inflammation over time were associated with effort-related food intake by fitting the same model while adding ΔINFLA-score (follow-up – baseline) to the interaction. If this ΔINFLA x Effort x Liking x Timepoint x Group interaction was not significant, we removed Group from the interaction and reported the ΔINFLA x Effort x Liking x Timepoint effects.

In all models, the outcome caloric intake was log-transformed to correct for skewed distribution of the model’s residuals. All models were adjusted for Age and baseline BMI and intervention models were additionally adjusted for INFLA-score at baseline. All models included a random intercept for participant but more complex random-effects structures were not included due to convergence issues.

Finally, we report mean+SD appetite scores, and liking, wanting and caloric intake per product, and tested its correlations with INFLA-score and ΔINFLA using partial Spearman correlations, adjusted for age, baseline BMI, and baseline INFLA-score.

### Inflammation and motivated behaviour in daily life measured with EMA

We included participants in the EMA analysis when they had at least 24 questionnaires (≈50% of max. number of questionnaires) completed. We categorized the type of activities reported as high effort (i.e. eating or drinking – self-prepared eating or drinking – other, exercising or moving, working or studying, doing groceries or shopping, or household tasks) or low effort (i.e. eating or drinking – takeaway or eating out, relaxing) activities.

First, we investigated the association between INFLA-score and motivation-related EMA variables: anticipation of the next activity (scale: 0-7), activity type (low-effort/high-effort), activity completion (yes/no), and subjective effort (scale: 0-7). These associations were tested using mixed linear and logistic regression modelling with each EMA variable as outcome and INFLA-score at the fixed factor of interest. We investigated the effect of the intervention on each EMA variable using mixed regression including Timepoint, Group, and their interaction as fixed factors of interest. Furthermore, we studied whether changes in inflammation were related to changes in EMA outcomes by fitting the same model while adding ΔINFLA-score (follow-up – baseline) to the Timepoint x Group interaction. When the ΔINFLA x Timepoint x Group interaction was not significant, the model was simplified by removing Group from the model, and the ΔINFLA x Timepoint effect was reported.

Second, we used mixed linear and logistic regression models to examine the time-lagged relationships between anticipation and activity completion as well as subjective effort of the activity. Across both the cross-sectional and intervention population, activity completion or subjective effort at time *t* were modelled as the outcome, with anticipation at the previous measurement (*t*–1) included as the predictor of interest.

Third, we examined whether INFLA-score was associated with these time-lagged relationships by including INFLA-score in interaction with anticipation (*t*-1) in these mixed-effects models. To test the effect of the intervention on these associations, we added an interaction between Timepoint and Group with anticipation (*t*-1). Finally, we tested whether changes in inflammation influenced these relationships, we added ΔINFLA-score to the Timepoint x Group x Anticipation (*t*-1) interaction. When this interaction was not significant, the model was simplified by removing Group from the model, and the ΔINFLA x Timepoint x x Anticipation (*t*-1) effect was reported.

According to our pre-registration (https://osf.io/2c3ax), we treated the analysis of the time-lagged associations between anticipation and activity completion as well as subjective effort as confirmatory. Corresponding p-values within each study were corrected for multiple comparisons using the Benjamini–Hochberg procedure. All mixed-effects models were corrected for age, BMI at baseline, and time since first beep, and included a random intercept for participant. Time-lagged models were additionally adjusted for the time-lagged version of the outcome. Intervention models were additionally corrected for INFLA-score at baseline and included a random slope for Timepoint.

### Inflammation and reported diet quality

To assess underreporting in the FFQ, we divided the total caloric intake as measured by the FFQ by the participants’ basal metabolic rate (EI:BMR) calculated by the Mifflin-St Jeor equation (Frankenfield, Roth-Yousey, and Compher 2005; Mifflin et al. 1990). Using a Physical Activity Level (PAL) of 1.55, the number of underreporters was defined as EI:BMR<1.1, following the Goldberg method (Black 2000).

Furthermore, due to skewed distribution of the DHD variables, we used partial Spearman correlations to assess the association between INFLA-score and DHD-guideline adherence, and daily caloric, macronutrient and micronutrient intake in the cross-sectional study. We tested the effect of the intervention using mixed linear regression modelling including Timepoint, Group, and their interaction as fixed factors of interest.

Furthermore, we tested how changes in INFLA-score were related to changes in DHD-guideline adherence and macro– and micronutrient intake by using partial Spearman correlations, adjusting for baseline outcome values.

All partial Spearman correlations and mixed models were adjusted for age, baseline BMI, and EI:BMR to correct for possible under– or overreporting. Mixed models and Spearman correlations assessing intervention effects were additionally adjusted for baseline INFLA-score.

### Inflammation and mood, reward sensitivity, and general health questionnaires

We examined the associations between INFLA-score and questionnaire-based measures of mood, reward sensitivity, and general health, as well as the effects of the intervention and changes in inflammation on these outcomes. Analyses were conducted using the same partial Spearman correlations and linear mixed regression models described above for diet-related outcomes.

### Relation between task-related effort aversion and ecological valid outcomes

Finally, we assessed how the lab-based outcome measures reported in our previous more mechanistic paper (Scholing et al., preprint) related to the more ecologically valid measures in the current paper. We related the task-derived behavioural derivatives and associated region-of-interest brain signal in the dorso-medial prefrontal cortex (dmPFC) to the effort-related intake as measured by the bogus taste test, the EMA outcomes, and the FFQ DHD intake. The effort aversion index reflects the acceptance rate of offers with increasing effort levels during a food-related effort-based decision-making task performed during functional MRI (Scholing et al. preprint, 2026). For interpretation purposes, we reversed the scores so that higher numbers indicate more effort aversion. The dmPFC region-of-interest (ROI) values reflect the effect of increasing effort level on brain signal in the a priori selected ROI during the offer presentation on screen.

We studied the relation of effort aversion and dmPFC signal with effort-related food intake by using the same mixed models as used to study the role of INFLA-score or dINFLA-score, but by replacing INFLA-score by effort aversion and dmPFC activity.

Furthermore, we studied the relation of effort aversion and dmPFC signal with EMA outcomes by performing similar regression analysis but by replacing INFLA-score with effort aversion and dmPFC signal.

Finally, we studied the relationship of effort aversion and dmPFC signal with diet quality by performing partial Spearman correlations between effort aversion and dmPFC signal and the FFQ outcomes. The correlations were adjusted for age and BMI.

## Results

### Demographics

We included 154 women in the cross-sectional study. Seven participants were subsequently excluded due to missing data on C-reactive protein (CRP), white blood cell count (WBC), platelets or neutrophil-to-lymphocyte ratio (NLR), and two due to abnormally high NLR values (>5), resulting in a sample of 145 participants. Of these, 144 completed the bogus taste test, 132 participants completed a sufficient number of forms (>=24) during the EMA, and 134 completed the food frequency questionnaire.

We included 59 participants in the randomized controlled trial, of whom 24 were included in the colchicine group and 25 in the placebo group. Two participants were excluded due to abnormally high NLR values (>5) at baseline. From these 57 participants, 56 completed the bogus taste test, 48 participants completed a sufficient number of forms (>=24) during the EMA, and 52 completed the food frequency questionnaire at both baseline and follow-up.

The demographic characteristics of all participants with data on any of the three outcomes are shown in Table 2. At baseline, the colchicine group had a higher NLR compared to the placebo group (mean±standard deviation (SD): 2.25±0.63 vs. 1.91±0.63, respectively, p = 0.045). The groups did not differ in any other baseline demographic, inflammatory or metabolic characteristics.

**Table 2:** Baseline characteristics of the cross-sectional and intervention study populations.

|  | Cross-sectional study<br>(n=145)<sup>a</sup> | Intervention study (n=57)<sup>a</sup> |  | P-value<br>(colchicine vs placebo)<sup>c</sup> |
| --- | --- | --- | --- | --- |
|  |  | Colchicine group<br>(n=26) | Placebo group<br>(n=31) |  |
|  | Mean ± SD or n (%) | Mean ± SD or n (%) | Mean ± SD or n (%) |  |
| Age (years)<sup>b</sup> | 47 (17) | 47 (16) | 47 (16) | 0.422 |
| BMI (kg/m<sup>2</sup>) | 35.5 ± 4.4 | 37.2 ± 3.7 | 38.6 ± 4.5 | 0.201 |
| Weight (kg) | 102 ± 15 | 107 ± 11 | 112 ± 13 | 0.109 |
| Waist-to-hip ratio | 0.90 ± 0.10 | 0.89 ± 0.06 | 0.90 ± 0.09 | 0.636 |
| Ethnicity (% Dutch) | 139 (96) | 26 (100) | 31 (100) | n.a. |
| Education (ISCED score) | 3.49 ± 1.35 | 3.31 ± 1.43 | 3.26 ± 1.15 | 0.689 |
| Hormonal birth control (%) | 49 (34) | 11 (42) | 12 (39) | 0.783 |
| Systolic Blood Pressure (mmHg) | 128 ± 15 | 133 ± 16 | 131 ± 12 | 0.618 |
| Diastolic Blood Pressure (mmHg) | 74 ± 10 | 76 ± 11 | 76 ± 10 | 0.893 |
| C-reactive protein (mg/L)<sup>b</sup> | 3.1 (5.0) | 6.0 (9.1) | 5.2 (4.6) | 0.689 |
| White blood cell count (*10<sup>3</sup>/mL) | 6.07 ± 1.36 | 6.76 ± 1.61 | 6.23 ± 1.62 | 0.230 |
| Neutrophil-lymphocyte ratio | 2.02 ± 0.66 | 2.25 ± 0.63 | 1.91 ± 0.61 | 0.045* |
| Neutrophil count (*10<sup>3</sup>/mL) | 3.52 ± 1.30 | 4.11 (1.47) | 3.57 (1.32) | 0.068 |
| Lymphocytes count (*10<sup>3</sup>/mL) | 1.82 ± 0.58 | 1.78 (0.79) | 1.98 (0.71) | 0.941 |
| Platelets count (*10<sup>3</sup>/mL) | 271±55 | 284 ± 63 | 273 ± 48 | 0.478 |
| INFLA score (SD) | N.a. | 0.08 ± 0.63 | -0.16 ± 0.49 | 0.108 |
| IL-6 (pg/mL)<sup>b</sup> | 1.25 (0.96) | 2.04 (0.92) | 1.83 (0.96) | 0.435 |
| Adiponectin (µg/mL) | 6.25 ± 3.44 | 5.27 ± 2.75 | 4.45 ± 2.27 | 0.232 |
| Leptin (ng/ml) | 94.5 ± 42.8 | 86.0 ± 36.3 | 98.6 ± 45.6 | 0.253 |
| HbA1c (mmol/mol) | 36.8 ± 3.7 | 37.4 ± 3.9 | 36.7 ± 3.8 | 0.500 |
| Insulin, fasting (mE/L)<sup>b</sup> | 13 (9) | 15 (10) | 17 (11) | 0.293 |

<sup>a</sup>Baseline data is reported for all subjects who have at data on at least one of the outcome measurements. <sup>b</sup>Median (IQR) is reported due to skewed distribution of the variable. <sup>c</sup>Difference between intervention groups at baseline was tested by independent t-test for normally distributed continuous variables, Wilcoxon-test for skewed continuous variables, and chi-square test for categorical variables. BMI, body mass index; HOMA-IR, Homeostasis Model Assessment of Insulin Resistance; IL, interleukin; ISCED, International Standard Classification of Education; POMS, Profile of Mood States. \*P<0.05.

## Cross-sectional results

### Inflammation and effort-related food intake

We used a bogus taste test to measure the participants’ effort-related food intake. As expected, high-effort products were associated with lower caloric intake compared to low-effort food items across both study samples (β –0.51 log(kcal) (95% CI –0.61, –0.42), R^2^_partial_ =0.056, p<0.001) (Fig. 2a). In addition, as expected, higher liking of the product was related to more caloric intake of these products (β 0.45 log(kcal) (95% CI 0.40, 0.50), R^2^_partial_ =0.153, p<0.001). Liking did not moderate the association between effort level and caloric intake (effortLevel x Liking: β –0.08 log(kcal) (95% CI –0.18, 0.02), R^2^_partial_ =0.001, p=0.121).

**Fig. 2.**
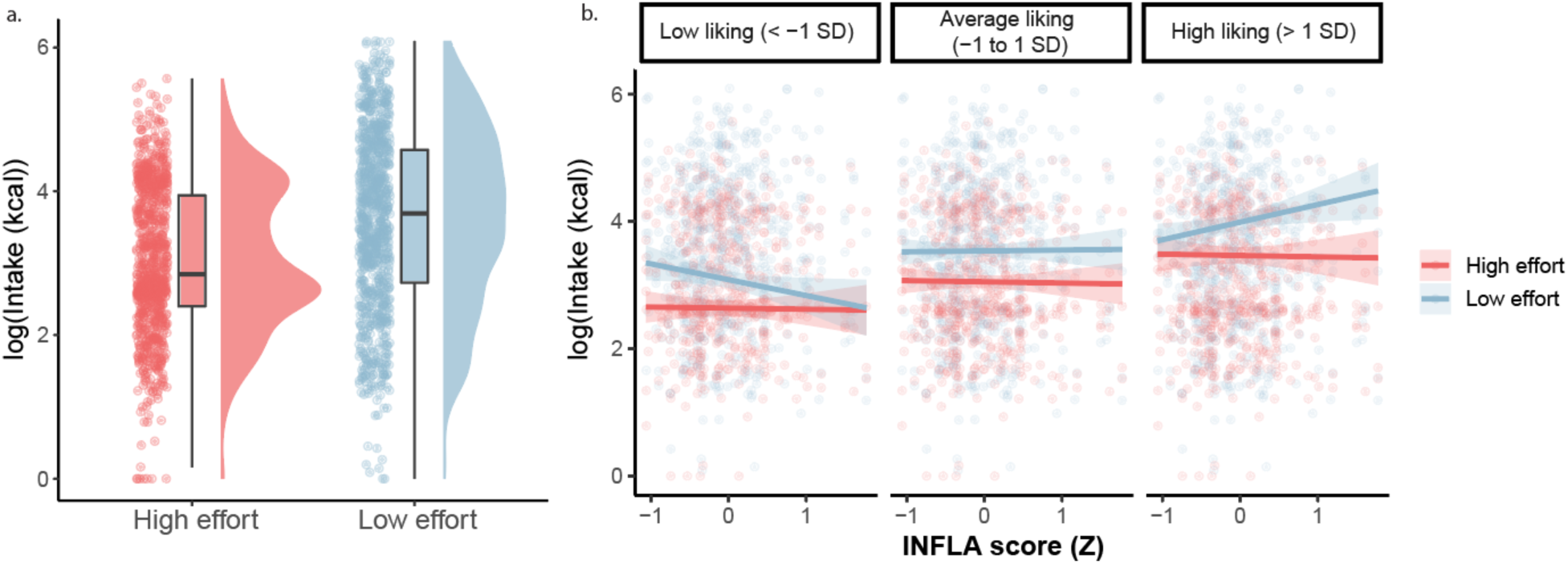
Effort-related food intake as measured by a bogus food taste test. Caloric food intake was log-transformed due to skewed distribution of the residuals. a. Caloric food intake for high– and low-effort food products. b. Interaction between INFLA score, effort, and liking predicting caloric food intake, as tested using linear mixed-effects modelling (β –0.2C SD (S5% CI – 0.47, –0.0C), R^2^_partial_ =0.00C, p = 0.011).

Furthermore, mixed linear regression analysis revealed that a higher INFLA score was related to lower caloric intake of high-effort food items, but only when they reported higher liking for the food item (INFLA x effortLevel x liking: β –0.26 SD (95% CI –0.47, –0.06), R^2^_partial_ =0.006, p=0.011) (Fig. 2b). In other words, participants with higher inflammation levels consumed more of the low-effort food items that they liked than participants with lower levels of inflammation. Full regression models can be found in Supplementary Table 3.

INFLA-score was related to higher hunger scores and more wanting to eat (r=0.20, p=0.031 and r=0.21, p=0.020, respectively), but was not related to wanting, liking or intake of any of the specific food products (all p>0.05). Mean+SD appetite, liking, wanting, and caloric intake per product and its correlations with INFLA-score can be found in Supplementary Tables 1 and 2.

### Inflammation and motivated behaviour in daily life

We used EMA to measure motivated behaviour in daily life. We first tested how INFLA-score was related to the EMA motivational components. Higher INFLA score was related to a lower proportion of high-effort compared to low-effort activities (OR: 0.72 (95% CI 0.62, 0.84), R^2^_partial_ =0.007, p<0.001, Fig. 3b). Furthermore, higher INFLA score was marginally related to more anticipation (β 0.14 (95% CI: –0.02, 0.30), R^2^_partial_ =0.006, p=0.085), and was not related to activity completion or subjective effort of the completed activity (p>0.05, Supplementary Table 4).

**Fig. 3.**
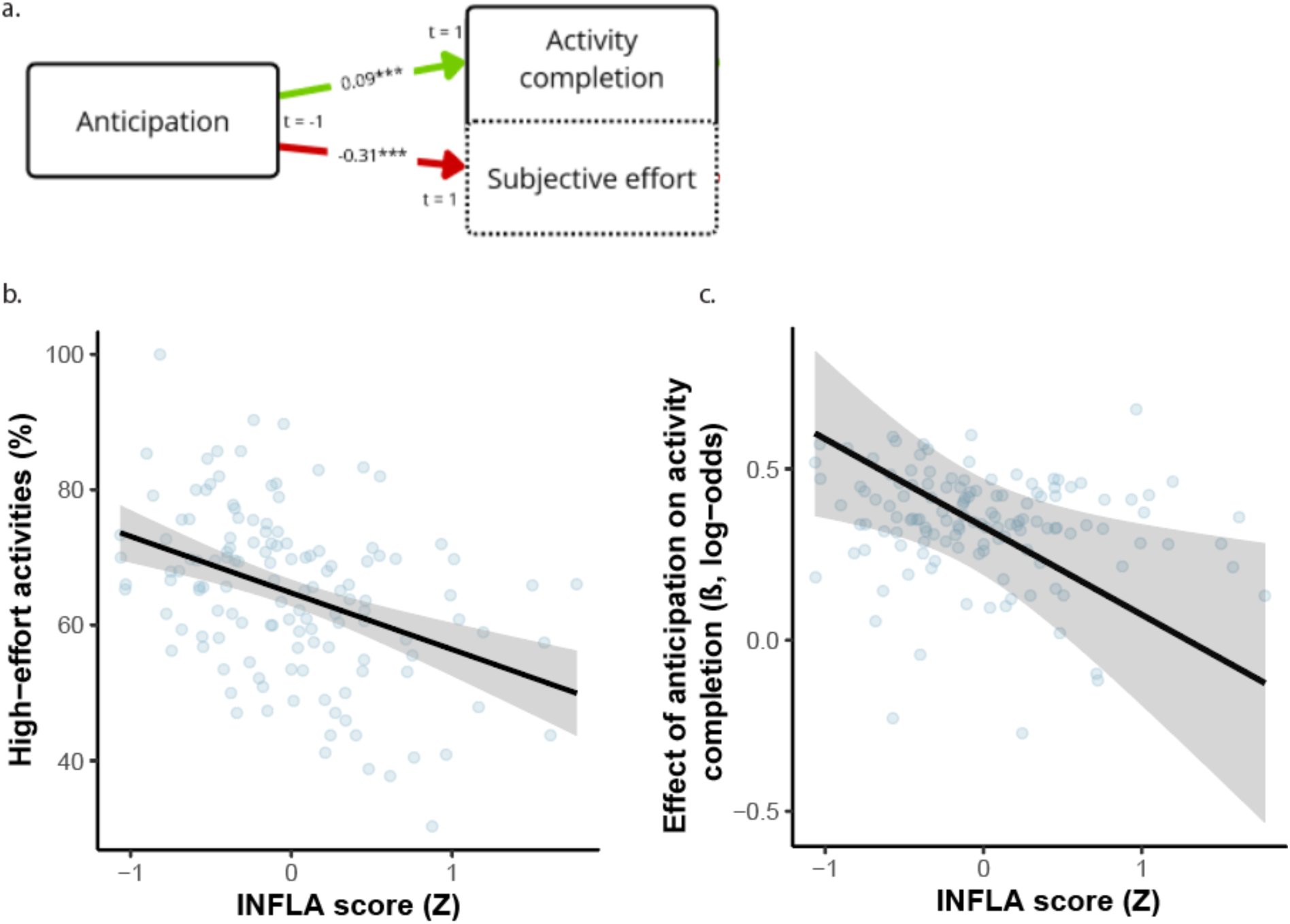
Results of the ecological momentary assessment (EMA). a. Schematic overview of motivational components including time-lagged associations between each other. Numbers represent model-derived partial correlation coefficients (R^2^_partial_), estimated using mixed linear regression models for the outcomes Anticipation and Subjective effort, and mixed logistic regression models for Activity completion. Green arrows indicate a positive relationship, whereas red arrows indicate a negative relationship. b. Association between INFLA-score and the percentage of high-effort activities completed, as tested by mixed logistic regression. c. Association between INFLA score and the relationship between anticipation and activity completion, as tested by mixed logistic regression. OR, odds ratio. *P<0.05, **P<0.01, ***P<0.001.

Second, we studied the time-lagged relationship between the motivational components (Fig. 3a, Table 3). More anticipation at the previous timepoint was related to a higher chance of completing the activity, (OR 1.30 (95% CI 1.19, 1.42), R^2^_partial_ =0.003, p<0.001). INFLA score negatively moderated this association between anticipation at the previous timepoint and activity completion (INFLA x Anticipation: OR 0.89 (95% CI: 0.80, 0.98), R^2^_partial_ =0.001, p_adjusted_=0.032; Fig. 3c), indicating that anticipation was less predictive of subsequent activity completion for participants with higher inflammation levels.

**Table 3:** Time-lagged associations between motivational components and the moderating role of inffammation. Coefficients were estimated using mixed linear regression models for continuous outcomes and mixed logistic regression models for dichotomous outcomes. All models are adjusted for the time-lagged version of the outcome variable, age, BMI at baseline, and time since the first beep. The models testing the effect of the intervention and change in inflammation are additionally adjusted for INFLA-score are baseline. All models included a random intercept for subject. Intervention models additionally included a random slope for Time. ^a^Odds ratios are reported for dichotomous outcome variables. ^b^Model additionally adjusted for the participant’s proportion of high-effort activities. *P<0.05, **P<0.01, ***P<0.001

| Predictor (time t-1) | Outcome (time t) | Beta/OR | 95% CI | R<sup>2</sup><sub>partial</sub> | P-value (uncorrected) | P-values (FDR-corrected) |
| --- | --- | --- | --- | --- | --- | --- |
| **Main behavioural associations** |  |  |  |  |  |  |
| Anticipation | Activity completion<sup>a</sup> (yes vs. no) | 1.15 | 1.09, 1.21 | 0.003 | <0.001*** | n.a. |
|  | Log(Subjective effort<sup>b</sup> (scale: 1-7)) | -0.09 | -0.10, -0.08 | 0.082 | <0.001*** | n.a. |
| **Moderation of INFLA** |  |  |  |  |  |  |
| Anticipation x INFLA | Activity completion<sup>a</sup> (yes vs. no) | 0.89 | 0.80, 0.98 | 0.001 | 0.016* | 0.032* |
|  | Log(Subjective effort<sup>b</sup> (scale: 1-7)) | <0.01 | -0.02, 0.01 | <0.001 | 0.741 | 0.741 |
| **Effect of colchicine vs. placebo** |  |  |  |  |  |  |
| Anticipation x Time x Group | Activity completion<sup>a</sup> (yes vs. no) | 1.08 | 0.85, 1.39 | <0.001 | 0.507 | 0.915 |
|  | Log(Subjective effort<sup>b</sup> (scale: 1-7)) | 0.01 | -0.03, 0.04 | <0.001 | 0.810 | 0.915 |
| **Effect of change in inflammation** |  |  |  |  |  |  |
| Anticipation x ΔINFLA x Time | Activity completion<sup>a</sup> (yes vs. no) | 0.02 | -0.30, 0.33 | <0.001 | 0.915 | 0.915 |
|  | Log(Subjective effort<sup>b</sup> (scale: 1-7)) | -0.02 | -0.08, 0.03 | <0.001 | 0.403 | 0.915 |

Furthermore, more anticipation at the previous timepoint was related to lower subjective effort ratings of the activity (log(β) –0.16 (95% CI: –0.18, –0.15), R^2^_partial_ =0.082, p=<0.001). INFLA-score did not moderate this association.

### Inflammation and diet quality

We used a FFQ to study their adherence to the Dutch Dietary Guidelines (DHD) by the Health Council of the Netherlands to assess overall diet quality in daily life. Based on the ratio of energy intake to BMR, 46% of participants in the cross-sectional study were classified as underreporters on the FFQ (EI:BMR<1.1).

Higher INFLA-score was associated with poorer adherence to the DHD-guidelines on legumes (partial Spearman ρ = –0.20, p=0.025), fruits (partial Spearman ρ = –0.17, p=0.047), and processed meat (partial Spearman ρ = –0.19, p=0.033) (Fig. 4a-c). Moreover, INFLA-score was related to lower intake of total iron (partial Spearman ρ = –0.24, p=0.005), non-heme iron (partial Spearman ρ = –0.26, p=0.003), and water (partial Spearman ρ = – 0.22, p=0.011). INFLA-score was not related to total DHD-adherence, any other DHD-components, macro-nutrient or micro-nutrient intake all p>0.1; Supplementary Table 5. Baseline diet quality and macro– and micronutrient intake can be found in Supplementary Table 6.

**Fig. 4.**
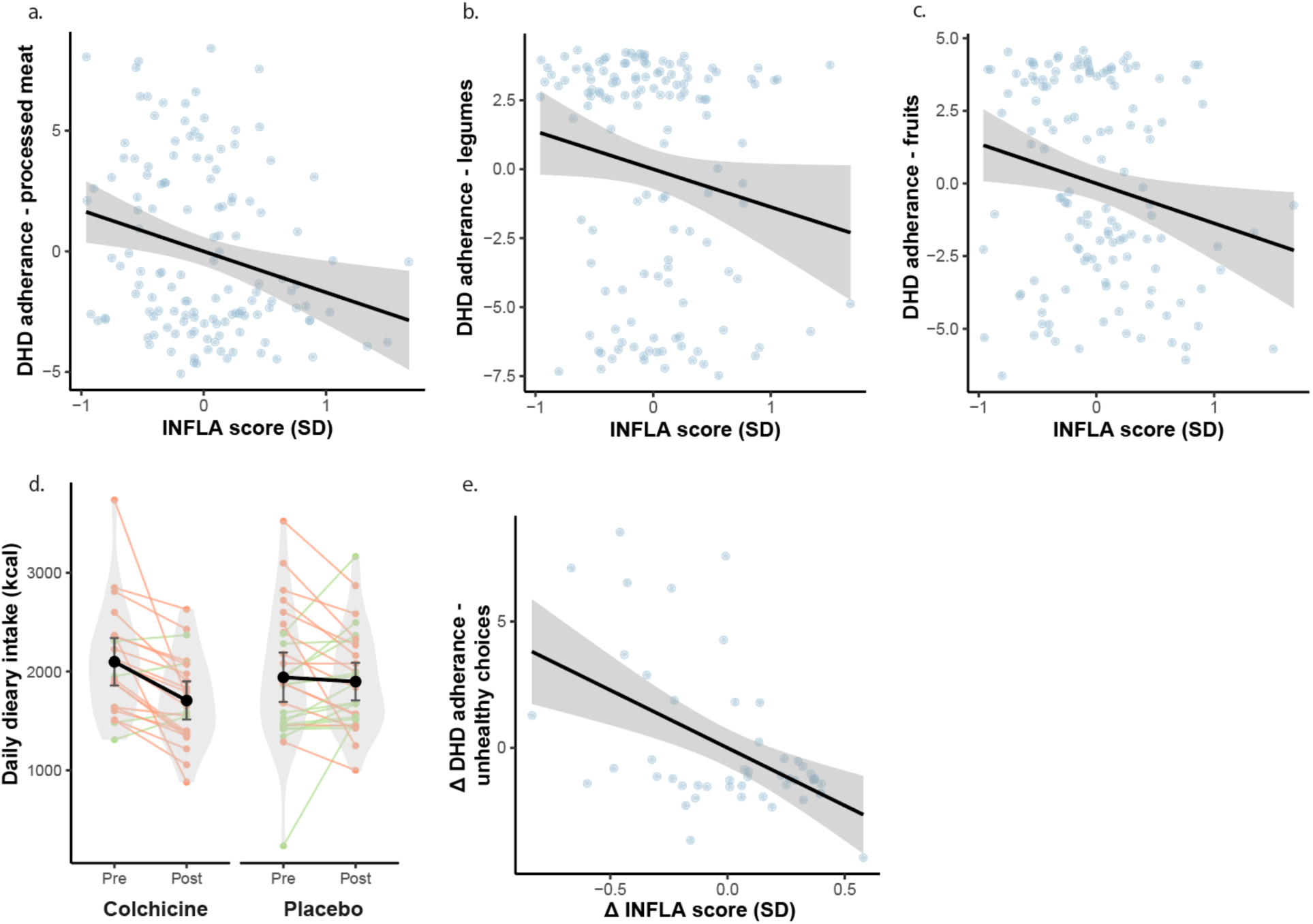
Associations between inffammation and the effect of colchicine on diet quality measured by the food frequency questionnaire. a, b, c. Association between INFLA-score and adherence to the Dutch Healthy Diet guidelines on processed meat, legumes and fruits in the cross-sectional study. Data points represent values adjusted for age and BMI. d. The effect of colchicine vs. placebo on daily caloric intake. e. The association between change in INFLA-score and change in adherence to the DHD guidelines on unhealthy choices. Data points represent values adjusted for age, baseline BMI and baseline INFLA-score. DHD, Dutch Healthy Diet; SD, standard deviation.

### Inflammation and mood, reward sensitivity, and general health

INFLA-score was associated with lower BAS score and lower scores on the BAS drive scores (β –0.19 (95% CI: –0.35 to –0.34), p=0.025; β –0.18 (95% CI: –0.34, –0.25), p=0.036). INFLA-score was not related to any of the other questionnaire outcomes (all p>0.1; Supplementary Table 7). Baseline questionnaire values can be found in Supplementary Table 8.

### Relation between task-related effort aversion and ecologically valid outcomes

More effort aversion during the effort-based decision-making task in the lab was related to less effort-related intake during the bogus taste test (EffortAversion x EffortLevel: β –0.16 (95% CI –0.28 to –0.03), p=0.018; Supplementary Table 9 and Supplementary Fig. 1). DmPFC signal was not related to effort-related food intake (Supplementary Table 10).

Task-related effort aversion and dmPFC signal were not related to any of the motivational components as measured by EMA (Supplementary Tables 11 and 12). Furthermore, task-related effort aversion and dmPFC signal were not related to DHD-adherence as measured by the FFQ (Supplementary Table 13 and 14).

## Intervention results

### Colchicine treatment reduced inflammation

Similar to our previous paper in approximately the same population (Scholing et al., preprint), we found that colchicine decreased inflammation compared to placebo, as indicated by the INFLA score (Time*Group: β=-0.10 SD, 95% CI: –0.15 to –0.04, p<0.001).

### Effect of intervention on effort-related food intake

We did not find a Time x Group intervention effect on effort-or liking-related food intake (Supplementary Table 3). In addition, a change in inflammation over time was not related to a change in effort– or liking-related food intake (all p>0.1; Supplementary Table 2).

Baseline, follow-up and intervention effects of appetite ratings, liking and food intake per food item are listed in Supplementary Table 1.

### Effect of intervention on motivated behaviour in daily life

We did not find any intervention effects on anticipation, activity completion, subjective effort of the completed activity, or the proportion of high-effort activities (Supplementary Table 4). We also did not find any moderating effects of the intervention on the time-lagged associations between anticipation, activity completion and subjective effort (Table 3).

### Effect of intervention on diet quality

Based on the ratio of energy intake to BMR, 44% of participants in the intervention study were classified as underreporters (EI:BMR<1.1).

Compared to placebo, colchicine treatment decreased daily caloric food intake (Time x Group: β –341 kcal (95% CI –620, –62), p=0.018; Fig. 4d). Exploratory analysis on macro– and micronutrient intake shows that colchicine decreased total protein intake, animal protein intake, total fat intake, saturated fat intake, mono-unsaturated food intake, trans fat intake, total carbohydrate intake, total iron intake, heme-iron intake, zinc intake and retinol equivalents intake (Supplementary Table 6).

In addition, a decrease in INFLA-score over time was related to better adherence to the guidelines on unhealthy food choices, i.e. more healthy food choices (partial Spearman ρ=-0.37, p=0.013; Fig. 4e), and better adherence to the guidelines on salt intake (partial Spearman ρ =-0.30, p=0.049). Change in INFLA score was not related to adherence to any of the other DHD subscales or altered macro– or micronutrient intake (Supplementary Table 5).

### Effect of intervention on mood, reward sensitivity, and general health

Colchicine treatment increased global sleep quality (1.57 (0.25, 2.89), p=0.021). Colchicine treatment did not affect any other questionnaire outcome, nor was change in INFLA-score related to any of these outcomes (Supplementary Table 7). Baseline and follow-up values of and intervention effects on all questionnaire outcomes can be found in Supplementary Table 8.

### Relation between change in task-related effort aversion and ecological valid outcomes

Intervention related change in task-related effort aversion and dmPFC signal were not related to effort-related food intake as measured by the bogus taste test, any of the motivational components as measured by the EMA, or DHD-adherence as measured by the FFQ (all p>0.1; Supplementary Tables 9-14).

## Discussion

We have previously demonstrated a causal role for inflammation in increasing effort aversion during food-related decision making in obesity in a laboratory setting (Scholing, et al. Preprint). We have now tested this principle in a more ecologically valid setting. We investigated the role of inflammation on food-related motivated behaviour in daily life in overweight and obesity using a multi-method approach combining a large cross-sectional study with a randomized controlled trial. In the cross-sectional study, we found that higher inflammation was related to lower intake of high-effort food items during a bogus taste test. Furthermore, higher inflammation related to less high-effort activities, and negatively moderated the relationship between anticipation and completion of the activity as measured by EMA. We did not find any effects of colchicine on these outcomes. Furthermore, we found that higher inflammation was associated with more intake of processed meat, and less intake of legumes, and fruits. Colchicine decreased overall daily caloric intake, and intake of saturated and mono-unsaturated fat, carbohydrates and (animal) protein. Finally, we found that a decrease in inflammation was related to better adherence to guidelines on healthy food choices. These results link inflammation as an important factor to food-related decisions and motivated behaviour in daily life in obesity.

Our findings in the cross-sectional study are in line with a growing body of evidence linking inflammation to effort aversion and lower motivated behaviour. Previously, we showed that obesity-related low-grade inflammation is associated with increased effort aversion, reflected by fewer high-effort choices during a food-related effort-based decision-making task in the MRI lab (Scholing, et al., 2025 preprint). Colchicine treatment reversed this effect by increasing high-effort food choices. This is also in line with a recent study from Chat et al. (2026) showing that more obesity-related inflammation was associated with lower high-effort choices in a monetary decision-making task in the lab (Chat et al. 2026). Similar effects have been reported in other populations. Treadway et al. (2025) showed that the anti-inflammatory drug infliximab decreased effort aversion in a depressed population with increased inflammation using a similar effort-based decision making task (Treadway et al. 2025). In healthy participants, LPS-induced inflammation also decreased high-effort choices during an effort-based decision making task (Draper et al. 2018). Together, these findings support a strong link between inflammation and effort aversion.

To date, the role of inflammation on effort aversion has largely been studied in well-controlled laboratory paradigms, which provide valuable insight into its underlying mechanisms. However, to fully understand its role in maladaptive eating behaviour and other types of motivated behaviour, it is important to translate these findings to everyday environments. Evidence from the role of inflammation on more ecologically valid measures is limited and relies on more indirect outcome measures, such as chronic low-grade inflammation causing lower physical activity in older adults, and less weight loss in bariatric patients (Hlebichuk et al. 2023; McLaughlin et al. 2024). By integrating inflammatory markers with effort-related measures in daily-life contexts, the present study provides initial evidence that low-grade inflammation may also relate to motivational processes and maladaptive eating behaviour beyond controlled experimental tasks.

More specifically, using a newly developed bogus taste test, we found that inflammation was related to lower intake of high-effort food items (i.e. food items requiring peeling or more chewing). In obesity, self-reported measures are particularly prone to underreporting (Braam et al. 1998; Santos-Báez et al. 2025), highlighting the importance for objective behavioural measures. Bogus taste tests provide such an alternative as they offer a more objective and behaviour-based assessment of actual food intake (Robinson et al. 2017). In line with this approach, we specifically designed such a taste test to assess effort-related food intake. Indeed, effort-related intake as measured by our newly developed taste test was associated with effort aversion in our more commonly used laboratory-based effort-based decision-making task. This supports the validity of this paradigm as an objective measure of effort-related food choice and indicates that this may be a useful tool for studying motivational processes underlying food choice, particularly in the context of inflammation.

However, we did not find any colchicine effects on food intake as measured by the bogus taste test. A limitation of bogus taste tests are that they are most effective when participants are unaware of the objective of the test, as more awareness has been associated with lower intake due to social desirability effects (Robinson et al. 2015). With repetition, the participants might become more aware of the purpose of the test, making the test less reliable for within-subject measurements (Robinson et al. 2017). This may have limited our ability to detect intervention-effects on this task.

Although we did not find colchicine effects on effort-related intake in the bogus taste test, we did find effects of colchicine on daily caloric intake as measured by the FFQ. Based on an exploratory analysis on macronutrient intake, the decrease in calories seemed to be related to lower intake of (animal) protein, saturated fats and carbohydrates. Together with our finding that a decrease in inflammation was related to better adherence to the Dutch Healthy Diet (DHD) guidelines on healthy choices, this indicates that the effect of colchicine on caloric intake might have been related to lower intake of high-caloric convenient foods. While no human studies have investigated causal effects of inflammation on dietary intake, these findings are in line with preclinical work showing that lowering diet-induced inflammation leads to lower caloric food intake (Alsaggar et al. 2020; Thornton et al. 2024; Wu et al. 2014; Yang, Li, and Wang 2024). For instance, pharmacological inhibition of the NLRP3 inflammasome—the same target we target using colchicine—decreased both food intake and body fat mass in mice with diet-induced obesity, with effect sizes comparable to the weight-loss medication semaglutide (Thornton et al. 2024). We extend these findings by providing evidence for a similar effect of inflammation on dietary intake in humans.

Notably, while we found effects of colchicine on caloric and specific macronutrient intake, we did not find effects on dietary patterns as measured by the DHD guidelines. This may reflect limited sensitivity of this type of composite diet quality scores to inflammation, potentially due to categorization and ceiling effects of such scores (Waijers, Feskens, and Ocké 2007). Given this, and the earlier-mentioned limitations of self-reported dietary measures in obesity, future studies should confirm the effect of inflammation on dietary patterns using alternative, more objective assessments of food intake.

Using EMA, we further show that inflammation is linked to motivational processes not only in the context of food, but also to general motivation in daily life. Higher levels of inflammation were related to reduced engagement in high-effort activities, in line with our findings on effort aversion in the lab (Scholing et al., preprint). In addition, inflammation was marginally linked to a weaker association between anticipation and activity completion, suggesting that individuals with higher inflammation were less likely to translate intentions into actual behaviour. Notably, a similar decoupling between anticipation and active behaviour has been observed with higher symptom severity in patients with depression (Bakker et al. 2017), indicating a potentially shared mechanism. Obesity and depression share a bidirectional relationship (Frank et al. 2022; Luppino et al. 2010; Martin-Rodriguez et al. 2016), in which inflammation acts as a mediating factor (Kaufmann et al. 2024; Milaneschi et al. 2019). In both conditions, inflammation has been related to weight loss difficulties (McLaughlin et al. 2024). Together, these findings therefore suggest that inflammation contributes to a transdiagnostic impairment in motivated behaviour, particularly on effort engagement and the translation of motivation into action, in both obesity and depression, which may partly explain difficulties with maintaining weight loss.

Similar to the bogus taste test, we did not find any effects of colchicine on these EMA outcomes. Ecologically valid measures of motivation have an inherently more heterogenous and multi-factorial nature than controlled laboratory tasks, as they are influenced by numerous contextual, behavioural, and environmental factors. In addition, day-to-day fluctuations in motivation drive may further increase variability in these outcomes (Hewitt et al. 2025). Furthermore, EMA may introduce assessment reactivity, where the repeated measurements increase the participants’ awareness of their behaviour, leading to an underestimation of typical behaviour (Wrzus and Neubauer 2023). This higher within– and between-subject variability may therefore have attenuated subtle pharmacological effects of colchicine on real-life motivated behaviour. Because the size of our study population was based on power calculations for the lab-controlled task rather than these more variable real-life measures, it is possibly that our design was insufficiently sensitive to detect modest effects of colchicine on these momentary measures of real-life behaviour. Furthermore, changing real-life motivated behaviour patterns may take longer than changes in laboratory-controlled tasks (Lally et al. 2010). Therefore, in the future, larger and longer intervention studies are needed to assess the causal effects of inflammation on daily motivation and effort-related food intake in obesity.

Another factor attenuating the effect of colchicine on the bogus taste test and EMA measures is the potentially bidirectional relationship between inflammation and motivated behaviour. While our earlier findings provide evidence that inflammation can increase effort aversion in laboratory-controlled tasks, in more ecologically valid settings reduced effortful behaviour might also contribute to increased inflammation. Especially poor diet quality, such as a higher intake of energy-dense or ultra-processed foods, often low-effort options, are known to cause low-grade inflammation (Grosso et al. 2022; Kopf et al. 2018; Tristan Asensi et al. 2023). Furthermore, reduced engagement in effortful activities, such as physical activity could promote sedentary behaviour, which has been associated with metabolic changes and systemic inflammation (Henson et al. 2013; Miao et al. 2024).

Consequently, our observational finding might not always translate into detectable effects in a short-term pharmacological intervention targeting inflammation alone. Nevertheless, the current colchicine’s effects on dietary intake measured with the FFQ does indicate that inflammation can causally affect eating behaviour in daily-life.

In conclusion, our findings show that obesity-related low-grade inflammation is associated with lower intake of high-effort food items, lower motivation for high-effort activities, and anticipation translating less often in activity engagement. Despite no evidence for causal effects on the bogus taste test and EMA (both known for their lower reliability with repeated measures), we could demonstrate that lowering inflammation decreases self-reported (unhealthy) food intake in daily life. This suggests that inflammation plays an important role in reduced effort-related motivation in obesity, which may bias food choices toward more convenient options and may compromise the ability to adhere to lifestyle changes. Future studies should confirm these placebo-controlled anti-inflammatory effects on daily food choice using more objective measures.

## Data Availability

All data and code needed to evaluate the conclusions of the present study will be available upon request to the authors.

## References

1. Alsaggar, Mohammad, Shifa Bdour, Qutaibah Ababneh, Tamam El-Elimat, Nidal Qinna, and Karem H. Alzoubi. 2020. “Silibinin Attenuates Adipose Tissue Inflammation and Reverses Obesity and Its Complications in Diet-Induced Obesity Model in Mice.” BMC pharmacology & toxicology 21(1): 8. doi:10.1186/s40360-020-0385-8.

2. AL-Suhaimi, Ebtesam A., and Adeeb Shehzad. 2013. “Leptin, Resistin and Visfatin: The Missing Link between Endocrine Metabolic Disorders and Immunity.” European Journal of Medical Research 18(1): 12. doi:10.1186/2047-783X-18-12.

3. Bakker, Jindra Myrthe, Liesbet Goossens, Iris Lange, Stijn Michielse, Koen Schruers, Ritsaert Lieverse, Machteld Marcelis, et al. 2017. “Real-Life Validation of Reduced Reward Processing in Emerging Adults with Depressive Symptoms.” Journal of Abnormal Psychology 126(6): 713–25. doi:10.1037/abn0000294.

4. Black, A. E. 2000. “Critical Evaluation of Energy Intake Using the Goldberg Cut-off for Energy Intake:Basal Metabolic Rate. A Practical Guide to Its Calculation, Use and Limitations.” International Journal of Obesity and Related Metabolic Disorders: Journal of the International Association for the Study of Obesity 24(9): 1119–30. doi:10.1038/sj.ijo.0801376.

5. Bonaccio, Marialaura, Augusto Di Castelnuovo, George Pounis, Amalia De Curtis, Simona Costanzo, Mariarosaria Persichillo, Chiara Cerletti, et al. 2016. “A Score of Low-Grade Inflammation and Risk of Mortality: Prospective Findings from the Moli-Sani Study.” Haematologica 101(11): 1434–41. doi:10.3324/haematol.2016.144055.

6. Braam, L. A., M. C. Ocké, H. B. Bueno-de-Mesquita, and J. C. Seidell. 1998. “Determinants of Obesity-Related Underreporting of Energy Intake.” American Journal of Epidemiology 147(11): 1081–86. doi:10.1093/oxfordjournals.aje.a009402.

7. Burgess, E., P. Hassmén, and K. L. Pumpa. 2017. “Determinants of Adherence to Lifestyle Intervention in Adults with Obesity: A Systematic Review.” Clinical Obesity 7(3): 123–35. doi:10.1111/cob.12183.

8. Buyken, Anette E., Janina Goletzke, Gesa Joslowski, Anna Felbick, Guo Cheng, Christian Herder, and Jennie C. Brand-Miller. 2014. “Association between Carbohydrate Quality and Inflammatory Markers: Systematic Review of Observational and Interventional Studies.” The American Journal of Clinical Nutrition 99(4): 813–33. doi:10.3945/ajcn.113.074252.

9. Buysse, D. J., C. F. Reynolds, T. H. Monk, S. R. Berman, and D. J. Kupfer. 1989. “The Pittsburgh Sleep Quality Index: A New Instrument for Psychiatric Practice and Research.” Psychiatry Research 28(2): 193–213. doi:10.1016/0165-1781(89)90047-4.

10. Carver, Charles S., and Teri L. White. 1994. “Behavioral Inhibition, Behavioral Activation, and Affective Responses to Impending Reward and Punishment: The BIS/BAS Scales.” Journal of Personality and Social Psychology 67(2): 319–33. doi:10.1037/0022-3514.67.2.319.

11. Chat, Iris Ka-Yi, Lina S Hansson, Mats Lekander, Charlotta Jacobsen, Sven Benson, Johannes Hebebrand, Vera Bender, et al. 2026. “Obesity, Low-Grade Inflammation, and Inflammatory Response to Immune Challenge Modulate Willingness to Expend Effort for Reward.” Brain, Behavior, and Immunity 136: 106548. doi:10.1016/j.bbi.2026.106548.

12. Chong, T.T.-J., V. Bonnelle, and M. Husain. 2016. “Quantifying Motivation with Effort-Based Decision-Making Paradigms in Health and Disease.” In Progress in Brain Research, Elsevier, 71–100. doi:10.1016/bs.pbr.2016.05.002.

13. Dantzer, R. 2001. “Cytokine-Induced Sickness Behavior: Mechanisms and Implications.” Annals of the New York Academy of Sciences 933: 222–34. doi:10.1111/j.1749-6632.2001.tb05827.x.

14. De Marco, Riccardo, Andrew W. Barritt, Mara Cercignani, Giulia Cabbai, Alessandro Colasanti, and Neil A. Harrison. 2023. “Inflammation-Induced Reorientation of Reward versus Punishment Sensitivity Is Attenuated by Minocycline.” Brain, Behavior, and Immunity 111: 320–27. doi:10.1016/j.bbi.2023.04.010.

15. Draper, Amelia, Rebecca M Koch, Jos WM van der Meer, Matthew AJ Apps, Peter Pickkers, Masud Husain, and Marieke E van der Schaaf. 2018. “Effort but Not Reward Sensitivity Is Altered by Acute Sickness Induced by Experimental Endotoxemia in Humans.” Neuropsychopharmacology 43(5): 1107–18. doi:10.1038/npp.2017.231.

16. Epstein, Leonard H., Jennifer L. Temple, Brad J. Neaderhiser, Robbert J. Salis, Richard W. Erbe, and John J. Leddy. 2007. “Food Reinforcement, the Dopamine D2 Receptor Genotype, and Energy Intake in Obese and Nonobese Humans.” Behavioral neuroscience 121(5): 877–86. doi:10.1037/0735-7044.121.5.877.

17. Eussen, Simone Jpm, Martien Cjm van Dongen, Nicole Eg Wijckmans, Saskia Meijboom, Henny Am Brants, Jeanne Hm de Vries, H. Bas Bueno-de-Mesquita, et al. 2018. “A National FFQ for the Netherlands (the FFQ-NL1.0): Development and Compatibility with Existing Dutch FFQs.” Public Health Nutrition 21(12): 2221–29. doi:10.1017/S1368980018000885.

18. Frank, Philipp, Markus Jokela, G. David Batty, Camille Lassale, Andrew Steptoe, and Mika Kivimäki. 2022. “Overweight, Obesity, and Individual Symptoms of Depression: A Multicohort Study with Replication in UK Biobank.” Brain, Behavior, and Immunity 105: 192–200. doi:10.1016/j.bbi.2022.07.009.

19. Frankenfield, David, Lori Roth-Yousey, and Charlene Compher. 2005. “Comparison of Predictive Equations for Resting Metabolic Rate in Healthy Nonobese and Obese Adults: A Systematic Review.” Journal of the American Dietetic Association 105(5): 775–89. doi:10.1016/j.jada.2005.02.005.

20. Giesen, Janneke C.A.H., Remco C. Havermans, Anne Douven, Mignon Tekelenburg, and Anita Jansen. 2010. “Will Work for Snack Food: The Association of BMI and Snack Reinforcement.” Obesity 18(5): 966–70. doi:10.1038/oby.2010.20.

21. Greenberg, Ilana, Meir J. Stampfer, Dan Schwarzfuchs, and Iris Shai. 2009. “Adherence and Success in Long-Term Weight Loss Diets: The Dietary Intervention Randomized Controlled Trial (DIRECT).” Journal of the American College of Nutrition 28(2): 159–68. doi:10.1080/07315724.2009.10719767.

22. Grosso, Giuseppe, Daniela Laudisio, Evelyn Frias-Toral, Luigi Barrea, Giovanna Muscogiuri, Silvia Savastano, and Annamaria Colao. 2022. “Anti-Inflammatory Nutrients and Obesity-Associated Metabolic-Inflammation: State of the Art and Future Direction.” Nutrients 14(6): 1137. doi:10.3390/nu14061137.

23. Henson, Joseph, Thomas Yates, Charlotte L. Edwardson, Kamlesh Khunti, Duncan Talbot, Laura J. Gray, Thomas M. Leigh, Patrice Carter, and Melanie J. Davies. 2013. “Sedentary Time and Markers of Chronic Low-Grade Inflammation in a High Risk Population.” PloS One 8(10): e78350. doi:10.1371/journal.pone.0078350.

24. Hewitt, Samuel R. C., Agnes Norbury, Quentin J. M. Huys, and Tobias U. Hauser. 2025. “Day-to-Day Fluctuations in Motivation Drive Effort-Based Decision-Making.” Proceedings of the National Academy of Sciences 122(12): e2417964122. doi:10.1073/pnas.2417964122.

25. Hlebichuk, Jeanne L., Randall J. Gretebeck, Mauricio Garnier-Villarreal, Linda B. Piacentine, Maharaj Singh, and Kimberlee A. Gretebeck. 2023. “Physical Activity, Inflammation, and Physical Function in Older Adults: Results From the Health & Retirement Study.” Biological Research for Nursing 25(1): 24–32. doi:10.1177/10998004221111217.

26. Kaufmann, Lisa-Katrin, Emma Custers, Debby Vreeken, Jessica Snabel, Martine C. Morrison, Robert Kleemann, Maximilian Wiesmann, et al. 2024. “Additive Effects of Depression and Obesity on Neural Correlates of Inhibitory Control.” Journal of Affective Disorders 362: 174–85. doi:10.1016/j.jad.2024.06.093.

27. Kirby, K. N., and N. N. Maraković. 1996. “Delay-Discounting Probabilistic Rewards: Rates Decrease as Amounts Increase.” Psychonomic Bulletin & Review 3(1): 100–104. doi:10.3758/BF03210748.

28. Kopf, Julianne C., Mallory J. Suhr, Jennifer Clarke, Seong-Il Eyun, Jean-Jack M. Riethoven, Amanda E. Ramer-Tait, and Devin J. Rose. 2018. “Role of Whole Grains versus Fruits and Vegetables in Reducing Subclinical Inflammation and Promoting Gastrointestinal Health in Individuals Affected by Overweight and Obesity: A Randomized Controlled Trial.” Nutrition Journal 17(1): 72. doi:10.1186/s12937-018-0381-7.

29. Lally, Phillippa, Cornelia H. M. van Jaarsveld, Henry W. W. Potts, and Jane Wardle. 2010. “How Are Habits Formed: Modelling Habit Formation in the Real World.” European Journal of Social Psychology 40(6): 998–1009. doi:10.1002/ejsp.674.

30. Lambregts, B. I. H. M., E. Vassena, A. Jansen, D. E. Stremmelaar, P. Pickkers, M. Kox, E. Aarts, and M. E. van der Schaaf. 2023. “Fatigue during Acute Systemic Inflammation Is Associated with Reduced Mental Effort Expenditure While Task Accuracy Is Preserved.” Brain, Behavior, and Immunity 112: 235–45. doi:10.1016/j.bbi.2023.05.013.

31. Lasselin, Julie, Michael T. Treadway, Tamara E. Lacourt, Anne Soop, Mats J. Olsson, Bianka Karshikoff, Sofie Paues-Göranson, et al. 2017. “Lipopolysaccharide Alters Motivated Behavior in a Monetary Reward Task: A Randomized Trial.” Neuropsychopharmacology: Official Publication of the American College of Neuropsychopharmacology 42(4): 801–10. doi:10.1038/npp.2016.191.

32. Lemstra, Mark, Yelena Bird, Chijioke Nwankwo, Marla Rogers, and John Moraros. 2016. “Weight Loss Intervention Adherence and Factors Promoting Adherence: A Meta-Analysis.” Patient Preference and Adherence 10: 1547–59. doi:10.2147/PPA.S103649.

33. Looman, Moniek, Edith JM Feskens, Mariëlle de Rijk, Saskia Meijboom, Sander Biesbroek, Elisabeth HM Temme, Jeanne de Vries, and Anouk Geelen. 2017. “Development and Evaluation of the Dutch Healthy Diet Index 2015.” Public Health Nutrition 20(13): 2289–99. doi:10.1017/S136898001700091X.

34. Lowe, Michael R., Meghan L. Butryn, Elizabeth R. Didie, Rachel A. Annunziato, J. Graham Thomas, Canice E. Crerand, Christopher N. Ochner, et al. 2009. “The Power of Food Scale. A New Measure of the Psychological Influence of the Food Environment.” Appetite 53(1): 114–18. doi:10.1016/j.appet.2009.05.016.

35. Luppino, Floriana S., Leonore M. de Wit, Paul F. Bouvy, Theo Stijnen, Pim Cuijpers, Brenda W. J. H. Penninx, and Frans G. Zitman. 2010. “Overweight, Obesity, and Depression: A Systematic Review and Meta-Analysis of Longitudinal Studies.” Archives of General Psychiatry 67(3): 220–29. doi:10.1001/archgenpsychiatry.2010.2.

36. Martin-Rodriguez, E., F. Guillen-Grima, E. Aubá, A. Martí, and A. Brugos-Larumbe. 2016. “Relationship between Body Mass Index and Depression in Women: A 7-Year Prospective Cohort Study. The APNA Study.” European Psychiatry: The Journal of the Association of European Psychiatrists 32: 55–60. doi:10.1016/j.eurpsy.2015.11.003.

37. Mata, Fernanda, Michael Treadway, Alastair Kwok, Helen Truby, Murat Yücel, Julie C. Stout, and Antonio Verdejo-Garcia. 2017. “Reduced Willingness to Expend Effort for Reward in Obesity: Link to Adherence to a 3-Month Weight Loss Intervention.” Obesity 25(10): 1676–81. doi:10.1002/oby.21948.

38. Mathar, D., Annette Horstmann, B. Pleger, A. Villringer, and J. Neumann. 2016. “Is It Worth the Effort? Novel Insights into Obesity-Associated Alterations in Cost-Benefit Decision-Making.” Frontiers in Behavioral Neuroscience 9. doi:10.3389/fnbeh.2015.00360.

39. McLaughlin, Anna P., Ellen Lambert, Rebecca Milton, Nicole Mariani, Melisa Kose, Naghmeh Nikkheslat, Olivia Patsalos, et al. 2024. “Peripheral Inflammation Associated with Depression and Reduced Weight Loss: A Longitudinal Study of Bariatric Patients.” Psychological Medicine 54(3): 601–10. doi:10.1017/S0033291723002283.

40. McNair, et al. 1971. Manual for the Profile of Mood States. San Diego, CA: Educational and Industrial Testing Service.

41. Miao, Shuchuan, Xiaoyan Wang, Lu Ma, and Chao You. 2024. “Sedentary Behavior from Television Watching Elevates GlycA Levels: A Bidirectional Mendelian Randomization Study.” PloS One 19(8): e0308301. doi:10.1371/journal.pone.0308301.

42. Mifflin, M. D., S. T. St Jeor, L. A. Hill, B. J. Scott, S. A. Daugherty, and Y. O. Koh. 1990. “A New Predictive Equation for Resting Energy Expenditure in Healthy Individuals.” The American Journal of Clinical Nutrition 51(2): 241–47. doi:10.1093/ajcn/51.2.241.

43. Milaneschi, Yuri, W. Kyle Simmons, Elisabeth F. C. van Rossum, and Brenda Wjh Penninx. 2019. “Depression and Obesity: Evidence of Shared Biological Mechanisms.” Molecular Psychiatry 24(1): 18–33. doi:10.1038/s41380-018-0017-5.

44. Miras, Alexander D., Robert N. Jackson, Sabrina N. Jackson, Anthony P. Goldstone, Torsten Olbers, Timothy Hackenberg, Alan C. Spector, and Carel W. le Roux. 2012. “Gastric Bypass Surgery for Obesity Decreases the Reward Value of a Sweet-Fat Stimulus as Assessed in a Progressive Ratio Task.” The American Journal of Clinical Nutrition 96(3): 467–73. doi:10.3945/ajcn.112.036921.

45. de Rijk, Mariëlle G., Anne I. Slotegraaf, Elske M. Brouwer-Brolsma, Corine W. M. Perenboom, Edith J. M. Feskens, and Jeanne H. M. de Vries. 2021. “Development and Evaluation of a Diet Quality Screener to Assess Adherence to the Dutch Food-Based Dietary Guidelines.” The British Journal of Nutrition 128(8): 1–11. doi:10.1017/S0007114521004499.

46. Robinson, Eric, Charlotte A. Hardman, Jason C. G. Halford, and Andrew Jones. 2015. “Eating under Observation: A Systematic Review and Meta-Analysis of the Effect That Heightened Awareness of Observation Has on Laboratory Measured Energy Intake.” The American Journal of Clinical Nutrition 102(2): 324–37. doi:10.3945/ajcn.115.111195.

47. Robinson, Eric, Ashleigh Haynes, Charlotte A. Hardman, Eva Kemps, Suzanne Higgs, and Andrew Jones. 2017. “The Bogus Taste Test: Validity as a Measure of Laboratory Food Intake.” Appetite 116: 223–31. doi:10.1016/j.appet.2017.05.002.

48. Rohm, Theresa V., Daniel T. Meier, Jerrold M. Olefsky, and Marc Y. Donath. 2022. “Inflammation in Obesity, Diabetes, and Related Disorders.” Immunity 55(1): 31–55. doi:10.1016/j.immuni.2021.12.013.

49. Rollins, Brandi Y., Eric Loken, Jennifer S. Savage, and Leann L. Birch. 2014. “Measurement of Food Reinforcement in Preschool Children. Associations with Food Intake, BMI, and Reward Sensitivity.” Appetite 72: 21–27. doi:10.1016/j.appet.2013.09.018.

50. Romieu, Isabelle, Laure Dossus, Simón Barquera, Hervé M. Blottière, Paul W. Franks, Marc Gunter, Nahla Hwalla, et al. 2017. “Energy Balance and Obesity: What Are the Main Drivers?” Cancer Causes & Control 28(3): 247–58. doi:10.1007/s10552-017-0869-z.

51. Saltiel, Alan R., and Jerrold M. Olefsky. 2017. “Inflammatory Mechanisms Linking Obesity and Metabolic Disease.” The Journal of Clinical Investigation 127(1): 1–4. doi:10.1172/JCI92035.

52. Santos-Báez, Leinys S., Michele N. Ravelli, Diana A. Díaz-Rizzolo, Collin J. Popp, Dympna Gallagher, Bin Cheng, Dale Schoeller, and Blandine Laferrère. 2025. “Dietary Misreporting: A Comparative Study of Recalls vs Energy Expenditure and Energy Intake by Doubly-Labeled Water in Older Adults with Overweight or Obesity.” BMC medical research methodology 25(1): 115. doi:10.1186/s12874-025-02568-4.

53. Shivappa, Nitin, Susan E. Steck, Thomas G. Hurley, James R. Hussey, and James R. Hébert. 2014. “Designing and Developing a Literature-Derived, Population-Based Dietary Inflammatory Index.” Public Health Nutrition 17(8): 1689–96. doi:10.1017/S1368980013002115.

54. Simon, Joe J., Johannes Zimmermann, Sheila A. Cordeiro, Ina Marée, David E. Gard, Hans-Christoph Friederich, Matthias Weisbrod, and Stefan Kaiser. 2018. “Psychometric Evaluation of the Temporal Experience of Pleasure Scale (TEPS) in a German Sample.” Psychiatry Research 260: 138–43. doi:10.1016/j.psychres.2017.11.060.

55. Singmann, Henrik, Ben Bolker, Jake Westfall, Frederik Aust, Mattan S. Ben-Shachar, Søren Højsgaard, John Fox, et al. 2025. “Afex: Analysis of Factorial Experiments.” https://cran.r-project.org/web/packages/afex/index.html (March 13, 2026).

56. Starkstein, S. E., H. S. Mayberg, T. J. Preziosi, P. Andrezejewski, R. Leiguarda, and R. G. Robinson. 1992. “Reliability, Validity, and Clinical Correlates of Apathy in Parkinson’s Disease.” The Journal of Neuropsychiatry and Clinical Neurosciences 4(2): 134–39. doi:10.1176/jnp.4.2.134.

57. Tchernof, André, and Jean-Pierre Després. 2013. “Pathophysiology of Human Visceral Obesity: An Update.” Physiological Reviews 93(1): 359–404. doi:10.1152/physrev.00033.2011.

58. “The Assessment of Binge Eating Severity among Obese Persons – PubMed.” https://pubmed.ncbi.nlm.nih.gov/7080884/ (October 3, 2025).

59. “The Assessment of Fatigue: Psychometric Qualities and Norms for the Checklist Individual Strength – PubMed.” https://pubmed.ncbi.nlm.nih.gov/28554371/ (October 3, 2025).

60. Thornton, Peter, Valérie Reader, Zsofia Digby, Pamela Smolak, Nicola Lindsay, David Harrison, Nick Clarke, and Alan P. Watt. 2024. “Reversal of High Fat Diet-Induced Obesity, Systemic Inflammation, and Astrogliosis by the NLRP3 Inflammasome Inhibitors NT-0249 and NT-0796.” The Journal of Pharmacology and Experimental Therapeutics 388(3): 813–26. doi:10.1124/jpet.123.002013.

61. Treadway, Michael T., Sarah M. Etuk, Jessica A. Cooper, Shabnam Hossein, Evan Hahn, Samantha A. Betters, Shiyin Liu, et al. 2025. “A Randomized Proof-of-Mechanism Trial of TNF Antagonism for Motivational Deficits and Related Corticostriatal Circuitry in Depressed Patients with High Inflammation.” Molecular Psychiatry 30(4): 1407–17. doi:10.1038/s41380-024-02751-x.

62. Tristan Asensi, Marta, Antonia Napoletano, Francesco Sofi, and Monica Dinu. 2023. “Low-Grade Inflammation and Ultra-Processed Foods Consumption: A Review.” Nutrients 15(6): 1546. doi:10.3390/nu15061546.

63. Waijers, Patricia M. C. M., Edith J. M. Feskens, and Marga C. Ocké. 2007. “A Critical Review of Predefined Diet Quality Scores.” British Journal of Nutrition 97(2): 219–31. doi:10.1017/S0007114507250421.

64. Ware, J. E., and C. D. Sherbourne. 1992. “The MOS 36-Item Short-Form Health Survey (SF-36). I. Conceptual Framework and Item Selection.” Medical Care 30(6): 473–83.

65. Wrzus, Cornelia, and Andreas B. Neubauer. 2023. “Ecological Momentary Assessment: A Meta-Analysis on Designs, Samples, and Compliance Across Research Fields.” Assessment 30(3): 825–46. doi:10.1177/10731911211067538.

66. Wu, Yizhen, Yinghua Yu, A. Szabo, Mei Han, and Xu-Feng Huang. 2014. “Central Inflammation and Leptin Resistance Are Attenuated by Ginsenoside Rb1 Treatment in Obese Mice Fed a High-Fat Diet.” PLoS ONE 9. doi:10.1371/journal.pone.0092618.

67. Yang, Jiaxin, Wanyi Li, and Yuanwei Wang. 2024. “Capsaicin Reduces Obesity by Reducing Chronic Low-Grade Inflammation.” International Journal of Molecular Sciences 25(16): 8979. doi:10.3390/ijms25168979.

68. Zhou, Heping, C. J. Urso, and Viren Jadeja. 2020. “<p>Saturated Fatty Acids in Obesity-Associated Inflammation</P>.” Journal of Inflammation Research 13: 1–14. doi:10.2147/JIR.S229691.

69. Zigmond, A. S., and R. P. Snaith. 1983. “The Hospital Anxiety and Depression Scale.” Acta Psychiatrica Scandinavica 67(6): 361–70. doi:10.1111/j.1600-0447.1983.tb09716.x.

