## Supplementary material for "Low-grade inflammation and daily life food-related motivation in obesity"

**Supplementary Table 1: Subjective appetite ratings and food intake of products in the bogus taste test**

| | Cross-sectional study (n=144) | Intervention study (n=56) | | | | | | |
|---|---|---|---|---|---|---|---|---|
| | | Colchicine (n=26) | | Placebo (n=30) | | | | |
| | | Baseline | Follow-up | Baseline | Follow-up | Time x Group Effect[b] | | |
| | Mean ± SD | Mean ± SD | Mean ± SD | Mean ± SD | Mean ± SD | β (95% CI) | $R^2_{partial}$ | P-value[a] |
| *General appetite (VAS, 0-100)* | | | | | | | | |
| Hunger | 65±20 | 71 ± 13 | 73 ± 17 | 68 ± 23 | 72 ± 18 | -1.8 (-11, 7.8) | 0.001 | 0.710 |
| Satiety[a] | 8 (3, 19) | 10 (3, 17) | 4 (0, 13) | 8 (3, 26) | 8 (2, 17) | -0.52 (-9.8, 8.8) | 0.000 | 0.911 |
| Thirst | 61 ± 23 | 60 ± 22 | 71 ± 17 | 63 ± 24 | 65 ± 21 | 9.2 (-3.1, 22) | 0.011 | 0.139 |
| Wanting to eat[a] | 75 (60, 88) | 75 (65, 85) | 79 (69, 93) | 79 (66, 87) | 81 (67, 89) | 0.61 (-8.2, 9.4) | 0.000 | 0.890 |
| Wanting to drink[a] | 77 (58, 90) | 79 (62, 91) | 88 (75, 95) | 82 (74, 92) | 78 (68, 90) | 12 (0.01, 24) | 0.020 | 0.050 |
| Wanting sweetness | 50 ± 26 | 51 ± 29 | 62 ± 27 | 48 ± 21 | 50 ± 26 | 7.8 (-4.4, 20) | 0.006 | 0.204 |
| Wanting savoury | 63 ± 24 | 68 ± 22 | 70 ± 20 | 64 ± 27 | 67 ± 26 | -1.6 (-15, 12) | 0.000 | 0.806 |
| *Wanting per product (VAS, 0-100)* | | | | | | | | |
| Tomatoes | 55 ± 28 | 52 ± 32 | 47 ± 30 | 54 ± 28 | 53 ± 32 | -6.1 (-16, 3.9) | 0.002 | 0.229 |
| Carrots | 51 ± 25 | 51 ± 34 | 48 ± 29 | 56 ± 19 | 60 ± 24 | -6.7 (-18, 42) | 0.004 | 0.223 |
| Pistachios[a] | 79 (58, 91) | 72 (40, 95) | 76 (64, 84) | 70 (51, 91) | 79 (64, 88) | 1.7 (-11, 14) | 0.000 | 0.789 |
| Cake | 59 ± 29 | 71 ± 21 | 63 ± 21 | 59 ± 25 | 57 ± 29 | -6.0 (-15, 3.1) | 0.004 | 0.189 |
| Cheese[a] | 69 (48, 87) | 72 (47, 88) | 69 (47, 83) | 72 (46, 88) | 76 (65, 84) | -6.6 (-17, 3.6) | 0.004 | 0.199 |
| Liquorice[a] | 35 (5, 63) | 34 (7, 62) | 24 (8, 64) | 28 (6, 62) | 28 (6, 56) | -1.5 (-13, 10) | 0.000 | 0.792 |
| Apple | 59 ± 25 | 66 ± 26 | 63 ± 20 | 58 ± 22 | 58 ± 24 | -2.6 (-14, 9.1) | 0.001 | 0.660 |
| Grapes | 69 ± 21 | 68 ± 15 | 72 ± 18 | 72 ± 21 | 72 ± 19 | 4.3 (-6.2, 15) | 0.003 | 0.417 |
| *Liking per product (VAS, 0-100)* | | | | | | | | |
| Tomatoes[a] | 64 ± 28 | 63 ± 29 | 57 ± 28 | 64 ± 29 | 57 ± 31 | -1.2 (-11, 8.2) | 0.000 | 0.803 |
| Carrots | 63 ± 24 | 63 ± 32 | 60 ± 28 | 67 ± 21 | 68 ± 24 | -3.1 (-11, 5.1) | 0.001 | 0.449 |
| Pistacchios[a] | 87 (70, 93) | 84 (49, 94) | 81 (66, 94) | 85 (68, 92) | 85 (78, 96) | 1.9 (-6.6, 10) | 0.001 | 0.658 |
| Cake[a] | 68 (50, 87) | 76 (65, 86) | 74 (57, 86) | 67 (57, 82) | 76 (61, 85) | -2.8 (-12, 6.1) | 0.001 | 0.529 |
| Cheese[a] | 79 (59, 91) | 81 (57, 92) | 74 (55, 95) | 83 (61, 90) | 80 (71, 83) | -4.1 (-13, 4.6) | 0.002 | 0.351 |
| Liquorice | 53 ± 30 | 53 ± 31 | 56 ± 29 | 51 ± 28 | 51 ± 28 | 2.2 (-7.9, 12) | 0.000 | 0.667 |
| Apple[a] | 74 (60, 87) | 80 (65, 91) | 71 (60, 78) | 68 (58, 86) | 72 (66, 85) | -9.8 (-17, 2.1) | 0.020 | 0.014* |
| Grapes[a] | 80 (68, 88) | 81 (67, 88) | 77 (62, 94) | 84 (74, 92) | 82 (69, 96) | -5.5 (-14, 2.9) | 0.008 | 0.197 |
| *Intake per product (kcal)* | | | | | | | | |
| Tomatoes[a] | 9 (4, 18) | 7 (4, 14) | 7 (3, 19) | 7 (4, 16) | 9 (4, 21) | -1.4 (-6.4, 3.5) | 0.001 | 0.564 |
| Carrots[a] | 8 (5, 13) | 10 (5, 19) | 7 (4, 17) | 9 (5, 16) | 10 (6, 17) | -3.7 (-6.9, -0.47) | 0.009 | 0.026* |
| Pistachios[a] | 32 (15, 62) | 39 (15, 62) | 50 (28, 81) | 27 (14, 70) | 43 (19, 85) | 19 (-13, 50) | 0.008 | 0.242 |
| Cake[a] | 69 (42, 135) | 76 (46, 138) | 119 (82, 164) | 57 (44, 91) | 105 (36, 174) | -22 (-75, 30) | 0.001 | 0.399 |
| Cheese[a] | 95 (56, 176) | 104 (41, 198) | 102 (66, 179) | 92 (58, 201) | 163 (86, 255) | -43 (-88, 1.4) | 0.011 | 0.057 |
| Liquorice[a] | 13 (12, 24) | 14 (12, 15) | 13 (12, 25) | 14 (13, 26) | 14 (13, 25) | -2.1 (-16, 11) | 0.001 | 0.755 |
| Apple[a] | 57 (20, 68) | 62 (41, 67) | 65 (28, 73) | 46 (13, 61) | 35 (6, 58) | 4.5 (-7.3, 16) | 0.002 | 0.444 |
| Grapes[a] | 24 (12, 39) | 20 (11, 39) | 26 (13, 52) | 32 (14, 48) | 47 (27, 61) | -5.6 (22, 11) | 0.003 | 0.501 |
| Total | 382 ± 183 | 410 ± 194 | 469 ± 198 | 344 ± 183 | 464 ± 239 | -44 (-136, 49) | 0.001 | 0.348 |

*[a]Median (IQR) is reported due to skewed distribution of the variable. [b]Effect of the intervention was tested by mixed linear regression modelling including Time and Group and their interaction as fixed factor of interest, age and baseline BMI as covariates and a random intercept for participant. BMI, body mass index; HOMA-IR, Homeostasis Model Assessment of Insulin Resistance; IL, interleukin; ISCED, International Standard Classification of Education; POMS, Profile of Mood States. *P<0.05.*

### Supplementary Table 2: Associations between (change in) inflammation and subjective appetite ratings and food intake of products in bogus taste test

| | *Cross-sectional study (n=144)* | | | *Intervention study (n=56)* | | |
|---|---|---|---|---|---|---|
| | *Correlations with INFLA-score* | | | *Correlations with ΔINFLA-score* | | |
| | Spearman's ρ | 95% CI | P-value | Spearman's ρ | 95% CI | P-value |
| *General appetite (VAS, 0-100)* | | | | | | |
| Hunger | 0.20 | 0.02, 0.36 | 0.031* | -0.16 | -0.45, 0.16 | 0.323 |
| Satiety[a] | -0.12 | -0.30, 0.05 | 0.172 | 0.28 | -0.04, 0.55 | 0.084 |
| Thirst | -0.05 | -0.22, 0.13 | 0.618 | 0.27 | -0.05, 0.54 | 0.098 |
| Wanting to eat[a] | 0.21 | 0.03, 0.37 | 0.020* | -0.12 | -0.42, 0.20 | 0.448 |
| Wanting to drink[a] | 0.07 | -0.11, 0.24 | 0.455 | 0.21 | -0.12, 0.49 | 0.205 |
| Wanting sweetness | -0.01 | -0.19, 0.17 | 0.933 | 0.08 | -0.24, 0.39 | 0.614 |
| Wanting savoury | 0.13 | -0.05, 0.30 | 0.158 | -0.16 | -0.45, 0.16 | 0.328 |
| *Wanting per product (VAS, 0-100)* | | | | | | |
| Tomatoes | -0.05 | -0.23, 0.13 | 0.585 | 0.03 | -0.29, 0.34 | 0.845 |
| Carrots | -0.13 | -0.30, 0.05 | 0.161 | 0.22 | -0.10, 0.50 | 0.171 |
| Pistachios[a] | -0.01 | -0.18, 0.17 | 0.941 | -0.15 | -0.44, 0.17 | 0.360 |
| Cake | 0.16 | -0.02, 0.33 | 0.085 | 0.35 | 0.04, 0.60 | 0.031* |
| Cheese[a] | 0.03 | -0.15, 0.20 | 0.779 | 0.001 | -0.31, 0.32 | 0.992 |
| Liquorice[a] | 0.11 | -0.06, 0.28 | 0.216 | -0.01 | -0.32, 0.31 | 0.955 |
| Apple | -0.06 | -0.24, 0.11 | 0.465 | 0.19 | -0.14, 0.47 | 0.257 |
| Grapes | -0.06 | -0.24, 0.12 | 0.509 | 0.05 | -0.27, 0.36 | 0.740 |
| *Liking per product (VAS, 0-100)* | | | | | | |
| Tomatoes[a] | -0.10 | -0.27, 0.08 | 0.270 | -0.04 | -0.35, 0.28 | 0.825 |
| Carrots | -0.06 | -0.24, 0.12 | 0.505 | 0.04 | -0.28, 0.35 | 0.816 |
| Pistacchios[a] | 0.06 | -0.12, 0.23 | 0.547 | -0.26 | -0.53, 0.06 | 0.107 |
| Cake[a] | 0.14 | -0.04, 0.31 | 0.122 | 0.29 | -0.03, 0.55 | 0.073 |
| Cheese[a] | 0.07 | -0.11, 0.25 | 0.424 | 0.14 | -0.18, 0.44 | 0.380 |
| Liquorice | 0.17 | -0.01, 0.34 | 0.056 | 0.03 | -0.29, 0.34 | 0.853 |
| Apple[a] | -0.10 | -0.27, 0.08 | 0.279 | 0.35 | 0.04, 0.60 | 0.030* |
| Grapes[a] | -0.07 | -0.24, 0.11 | 0.454 | 0.13 | -0.19, 0.43 | 0.416 |
| *Intake per product (kcal)* | | | | | | |
| Tomatoes[a] | -0.003 | -0.18, 0.17 | 0.974 | -0.21 | -0.49, 0.11 | 0.198 |
| Carrots[a] | 0.02 | -0.15, 0.20 | 0.790 | -0.01 | -0.32, 0.31 | 0.976 |
| Pistachios[a] | 0.13 | -0.05, 0.30 | 0.168 | 0.03 | -0.29, 0.34 | 0.872 |
| Cake[a] | 0.04 | -0.14, 0.21 | 0.675 | -0.09 | -0.39, 0.24 | 0.603 |
| Cheese[a] | 0.12 | -0.06, 0.29 | 0.179 | -0.07 | -0.37, 0.26 | 0.692 |
| Liquorice[a] | -0.01 | -0.19, 0.17 | 0.931 | 0.18 | -0.14, 0.47 | 0.264 |
| Apple[a] | -0.11 | -0.29, 0.06 | 0.208 | -0.21 | -0.49, 0.12 | 0.204 |
| Grapes[a] | 0.12 | -0.06, 0.29 | 0.186 | 0.26 | -0.06, 0.53 | 0.114 |
| Total | 0.09 | -0.09, 0.26 | 0.337 | 0.08 | -0.24, 0.38 | 0.614 |

*Partial spearman correlations were used to test the association between INFLA-score and ΔINFLA-score (follow-up – baseline) and appetite, liking, wanting and caloric intake scores. [a]Median (IQR) is reported due to skewed distribution of the variable. CI, 95% confidence interval; VAS, visual analogue scale. *P<0.05*

**Supplementary Table 3: Mixed linear regression models of the relationship between (change in) inflammation and effort-related caloric food intake as measured by the bogus taste test**

| | Model 1: Main task effects | | | Model 2: INFLA-score | | | Model 3: Group x Time | | | Model 4: ΔINFLA x Time | | |
|---|---|---|---|---|---|---|---|---|---|---|---|---|
| **Predictor** | **β (95% CI)** | **$R^2_{partial}$[a]** | **p-value** | **β (95% CI)** | **$R^2_{partial}$[a]** | **p-value** | **β (95% CI)** | **$R^2_{partial}$[a]** | **p-value** | **β (95% CI)** | **$R^2_{partial}$[a]** | **p-value** |
| Effort (high vs. low) | -0.51 (-0.61, -0.42) | 0.056 | <0.001*** | -0.49 (-0.60, -0.37) | 0.053 | <0.001*** | -0.52 (-0.66, -0.39) | 0.149 | <0.001*** | -0.56 (-0.70, -0.41) | 0.057 | <0.001*** |
| Liking (SD) | 0.45 (0.40, 0.50) | 0.153 | <0.001*** | 0.43 (0.37, 0.49) | 0.148 | <0.001*** | 0.46 (0.39, 0.53) | 0.055 | <0.001*** | 0.45 (0.37, 0.53) | 0.128 | <0.001*** |
| Age (SD) | -0.02 (-0.09, 0.05) | <0.001 | 0.575 | -0.04 (-0.11, 0.04) | 0.001 | 0.342 | -0.06 (-0.19, 0.07) | 0.003 | 0.343 | -0.07 (-0.20, 0.06) | 0.004 | 0.300 |
| BMI_t1 (SD) | -0.01 (-0.08, 0.05) | <0.001 | 0.669 | 0.01 (-0.07, 0.08) | <0.001 | 0.921 | -0.06 (-0.19, 0.07) | 0.003 | 0.336 | -0.04 (-0.17, 0.09) | 0.002 | 0.494 |
| INFLA_t1 (SD) | - | - | - | 0.00 (-0.14, 0.13) | <0.001 | 0.969 | -0.16 (-0.40, 0.07) | 0.007 | 0.168 | -0.16 (-0.43, 0.11) | 0.005 | 0.240 |
| ΔINFLA (SD) | - | - | - | - | - | - | - | - | - | 0.03 (-0.34, 0.41) | <0.001 | 0.859 |
| Group (Colchicine, Placebo) | - | - | - | - | - | - | <0.01 (-0.26, 0.27) | <0.001 | 0.974 | - | - | - |
| Time (t1, t2) | - | - | - | - | - | - | 0.16 (0.02, 0.30) | 0.005 | 0.022* | 0.16 (0.01, 0.31) | 0.005 | 0.034* |
| Effort x Liking | 0.08 (-0.18, 0.02) | 0.001 | 0.121 | -0.04 (-0.16, 0.08) | <0.001 | 0.529 | -0.13 (-0.28, 0.01) | 0.004 | 0.063 | -0.13 (-0.28, 0.03) | 0.003 | 0.102 |
| Group x Liking | - | - | - | - | - | - | 0.11 (-0.03, 0.26) | 0.003 | 0.132 | - | - | - |
| Group x Effort | - | - | - | - | - | - | 0.21 (-0.06, 0.48) | 0.002 | 0.133 | - | - | - |
| Time x Liking | - | - | - | - | - | - | 0.13 (-0.01, 0.27) | 0.004 | 0.066 | 0.12 (-0.03, 0.28) | 0.003 | 0.110 |
| Time x Effort | - | - | - | - | - | - | -0.21 (-0.48, 0.06) | 0.002 | 0.125 | -0.23 (-0.52, 0.06) | 0.003 | 0.124 |
| Time x Group | - | - | - | - | - | - | -0.03 (-0.30, 0.25) | <0.001 | 0.854 | - | - | - |
| INFLA_t1 x Effort | - | - | - | -0.03 (-0.24, 0.17) | <0.001 | 0.756 | - | - | - | - | - | - |
| INFLA_t1 x Liking | - | - | - | 0.13 (0.03, 0.23) | 0.005 | 0.013* | - | - | - | - | - | - |
| ΔINFLA x Effort | - | - | - | - | - | - | - | - | - | -0.24 (-0.59, 0.11) | 0.002 | 0.175 |
| ΔINFLA x Liking | - | - | - | - | - | - | - | - | - | -0.04 (-0.22, 0.13) | <0.001 | 0.628 |
| ΔINFLA x Time | - | - | - | - | - | - | - | - | - | 0.05 (-0.30, 0.40) | <0.001 | 0.772 |
| Time x Group x Effort | - | - | - | - | - | - | 0.09 (-0.45, 0.64) | <0.001 | 0.740 | - | - | - |
| Effort x Liking x Group | - | - | - | - | - | - | 0.07 (-0.21, 0.36) | <0.001 | 0.615 | - | - | - |
| Liking x Effort x Time | - | - | - | - | - | - | 0.06 (-0.22, 0.33) | <0.001 | 0.684 | 0.04 (-0.26, 0.35) | | 0.782 |
| Time x ΔINFLA x Effort | - | - | - | - | - | - | - | - | - | -0.15 (-0.84, 0.55) | <0.001 | 0.682 |
| Time x Group x Liking | - | - | - | - | - | - | 0.05 (-0.23, 0.33) | <0.001 | 0.725 | - | - | - |

| | | | | | | | | | | | | |
|---|---|---|---|---|---|---|---|---|---|---|---|---|
| Time x ΔINFLA x Liking | - | - | - | - | - | - | - | - | - | 0.04 (-0.30, 0.38) | <0.001 | 0.800 |
| Effort x Liking x ΔINFLA | - | - | - | - | - | - | - | - | - | -0.03 (-0.38, 0.32) | <0.001 | 0.872 |
| INFLA_t1 x Effort x Liking | - | - | - | -0.26 (-0.47, -0.06) | 0.006 | 0.011* | - | - | - | - | - | - |
| Time x Group x Effort x Liking | - | - | - | - | - | - | 0.07 (-0.49, 0.62) | <0.001 | 0.816 | - | - | - |
| Time x ΔINFLA x Effort x Liking | - | - | - | - | - | - | - | - | - | -0.15 (-0.83, 0.53) | <0.001 | 0.660 |

*Model 1 shows the results of the mixed linear regression model investigating the effect of effort and liking on caloric food intake in baseline data of all participants included in the cross-sectional and the intervention study. Model 2 shows the results of the linear regression model investigating the association between INFLA-score and effort- and liking-related food intake in the cross-sectional study. Model 3 shows the results of the linear regression model investigating the effects of colchicine on effort- and liking related food intake in the intervention study. Model 4 shows the results of the mixed linear regression model investigating the association between change in INFLA-score and change in effort- and liking-related food intake. In all models, the outcome Caloric Intake was log-transformed to correct of skewed distribution of the residuals. All continuous variables in the model were standardized. CI, confidence interval; SD, standard deviation.*

### Supplementary Table 4: Associations between (change in) inflammation and EMA outcomes, and the effect of colchicine on EMA outcomes

| Predictors (time *t-1*) | Outcome (time *t*) | β / OR | 95% CI | $R^2_{partial}$ | P-value |
|---|---|---|---|---|---|
| ***Associations with inflammation*** | | | | | |
| INFLA | Anticipation (scale: 1-7) | 0.14 | -0.02, 0.30 | 0.006 | 0.085 |
| INFLA | Activity completion (yes vs. no) | 1.11[a] | 0.73, 1.73 | <0.001 | 0.640 |
| INFLA | Subjective effort[b] (scale: 1-7) | -0.06 | -0.37, 0.25 | <0.001 | 0.698 |
| INFLA | Activity type (high vs. low effort) | 0.72[a] | 0.62, 0.84 | 0.007 | <0.001*** |
| ***Effect of intervention*** | | | | | |
| Group x Time | Anticipation (scale: 1-7) | 0.02 | -0.03, 0.08 | <0.001 | 0.428 |
| Group x Time | Activity completion (yes vs. no) | 1.00[a] | 0.85, 1.16 | <0.001 | 0.973 |
| Group x Time | Subjective effort[b] (scale: 1-7) | 0.01 | -0.07, 0.10 | <0.001 | 0.781 |
| Group x Time | Activity type (high vs. low effort) | 0.98[a] | 0.90, 1.05 | <0.001 | 0.545 |
| ***Change in inflammation*** | | | | | |
| ΔINFLA x Time | Anticipation (scale: 1-7) | 0.00 | -0.15, 0.14 | <0.001 | 0.945 |
| ΔINFLA x Time | Activity completion (yes vs. no) | 0.93[a] | 0.64, 1.35 | <0.001 | 0.731 |
| ΔINFLA x Time | Subjective effort[b] (scale: 1-7) | -0.06 | -0.20, 0.08 | <0.001 | 0.390 |
| ΔINFLA x Time | Activity type (high vs. low effort) | 1.01[a] | 0.83, 1.22 | <0.001 | 0.956 |
| **Change in inflammation x Group** | | | | | |
| ΔINFLA x Time x Group | Anticipation (scale: 1-7) | 0.09 | -0.11, 0.28 | 0.001 | 0.370 |
| ΔINFLA x Time x Group | Activity completion (yes vs. no) | 0.91[a] | 0.52, 1.62 | <0.001 | 0.757 |
| ΔINFLA x Time x Group | Subjective effort[b] (scale: 1-7) | -0.09 | -0.28, 0.10 | <0.001 | 0.334 |
| ΔINFLA x Time x Group | Activity type (high vs. low effort) | 1.34[a] | 0.79, 1.34 | <0.001 | 0.850 |

*Coefficients were estimated using mixed linear regression models for continuous outcomes and mixed logistic regression models for dichotomous outcomes. All models are adjusted for age, BMI at baseline, and time since the first beep. The models testing the effect of the intervention and change in inflammation are additionally adjusted for INFLA-score are baseline. All models included a random intercept for subject. Intervention models additionally included a random slope for Time.*

*[a]Odds ratios are reported for dichotomous outcome variables. [b]Model additionally adjusted for the participant's proportion of high-effort activities. [c]This model only included a random intercept for Participant and no random slope for Time due to convergency issues. ***P<0.001*

### Supplementary Table 5: Correlations between (change in) inflammation and diet quality and macro- and micronutrient intake

| | *Cross-sectional study (n=134), associations with INFLA* | | | *Intervention study (n=49), associations with ΔINFLA* | | |
|---|---|---|---|---|---|---|
| | **Spearman's ρ** | **95% CI** | **P-value** | **Spearman's ρ** | **95% CI** | **P-value** |
| ***DHD-score*** | | | | | | |
| Total | -0.13 | -0.3, 0.04 | 0.128 | -0.04 | -0.32, 0.24 | 0.799 |
| Vegetables | -0.06 | -0.23, 0.11 | 0.494 | 0.26 | -0.03, 0.5 | 0.092 |
| Fruits | -0.17 | -0.34, -0. | 0.047* | 0.01 | -0.27, 0.29 | 0.937 |
| Whole grain products | -0.14 | -0.31, 0.03 | 0.103 | 0.17 | -0.11, 0.43 | 0.259 |
| Legumes | -0.20 | -0.36, -0.03 | 0.025* | 0.17 | -0.11, 0.43 | 0.258 |
| Nuts | -0.05 | -0.22, 0.12 | 0.584 | -0.04 | -0.32, 0.24 | 0.800 |
| Dairy | -0.04 | -0.21, 0.14 | 0.679 | -0.04 | -0.32, 0.24 | 0.792 |
| Fish | -0.08 | -0.24, 0.1 | 0.385 | 0.08 | -0.21, 0.35 | 0.605 |
| Tea | 0.00 | -0.18, 0.17 | 0.964 | -0.07 | -0.34, 0.22 | 0.662 |
| Fats and oils | 0.10 | -0.08, 0.26 | 0.279 | -0.02 | -0.3, 0.26 | 0.910 |
| Coffee | 0.06 | -0.11, 0.23 | 0.502 | 0.12 | -0.17, 0.39 | 0.435 |
| Red meat | 0.05 | -0.13, 0.22 | 0.597 | 0.09 | -0.2, 0.36 | 0.557 |
| Processed meat | -0.19 | -0.35, -0.02 | 0.033* | -0.10 | -0.37, 0.19 | 0.522 |
| Sugar-sweetened beverages | 0.01 | -0.16, 0.18 | 0.923 | 0.05 | -0.23, 0.33 | 0.731 |
| Alcohol | 0.15 | -0.02, 0.31 | 0.084 | 0.16 | -0.13, 0.42 | 0.311 |
| Salt | -0.04 | -0.21, 0.13 | 0.624 | -0.30 | -0.53, -0.02 | 0.049* |
| Unhealthy choices | 0.02 | -0.15, 0.2 | 0.778 | -0.37 | -0.59, -0.1 | 0.013* |
| ***Nutrient intake*** | | | | | | |
| Total energy intake (kcal/d) | 0.14 | -0.02, 0.31 | 0.090 | 0.12 | -0.17, 0.38 | 0.454 |
| Protein total (g/d) | 0.05 | -0.13, 0.21 | 0.609 | 0.10 | -0.18, 0.37 | 0.507 |
| Plant protein (g/d) | -0.06 | -0.23, 0.11 | 0.473 | 0.19 | -0.1, 0.45 | 0.216 |
| Animal protein (g/d) | 0.10 | -0.07, 0.27 | 0.238 | 0.06 | -0.23, 0.33 | 0.704 |
| Fat total (g/d) | -0.03 | -0.2, 0.14 | 0.716 | 0.17 | -0.11, 0.43 | 0.257 |
| Saturated fat (gd) | 0.07 | -0.11, 0.24 | 0.443 | 0.19 | -0.1, 0.45 | 0.217 |
| Mono-unsaturated fat (g/d) | -0.04 | -0.21, 0.14 | 0.684 | 0.17 | -0.12, 0.43 | 0.283 |
| Poly-unsaturated (g/d) | -0.10 | -0.26, 0.08 | 0.278 | 0.13 | -0.16, 0.4 | 0.402 |
| Trans fat (g/d) | 0.16 | -0.01, 0.32 | 0.069 | 0.13 | -0.16, 0.39 | 0.410 |
| Linoleic acid (g/d) | -0.07 | -0.24, 0.1 | 0.435 | 0.09 | -0.19, 0.36 | 0.549 |
| ALA (g/d) | -0.07 | -0.23, 0.11 | 0.457 | 0.09 | -0.2, 0.36 | 0.571 |
| EPA (mg/d) | -0.12 | -0.29, 0.05 | 0.172 | 0.02 | -0.27, 0.3 | 0.914 |
| DHA (g/d) | -0.08 | -0.24, 0.1 | 0.387 | 0.00 | -0.28, 0.28 | 0.985 |
| Cholesterol (g/d) | -0.08 | -0.25, 0.09 | 0.364 | 0.07 | -0.21, 0.35 | 0.647 |
| Carbohydrates total (g/d) | 0.00 | -0.17, 0.17 | 0.977 | 0.19 | -0.1, 0.45 | 0.219 |
| Sugars (g/d) | -0.14 | -0.3, 0.04 | 0.124 | 0.15 | -0.14, 0.41 | 0.342 |
| Polysaccharides (g/d) | 0.14 | -0.03, 0.31 | 0.101 | 0.15 | -0.13, 0.42 | 0.321 |
| Fiber (g/d) | -0.11 | -0.27, 0.07 | 0.223 | 0.20 | -0.09, 0.46 | 0.192 |
| Water (ml/d) | -0.22 | -0.38, -0.05 | 0.011* | 0.18 | -0.11, 0.43 | 0.256 |
| Alcohol (g/d) | 0.15 | -0.02, 0.31 | 0.084 | -0.07 | -0.35, 0.21 | 0.629 |
| Calcium (mg/d) | -0.03 | -0.2, 0.15 | 0.768 | 0.09 | -0.19, 0.36 | 0.548 |
| Iron total (mg/d) | -0.24 | -0.4, -0.08 | 0.005** | 0.19 | -0.09, 0.45 | 0.209 |
| Heme iron (mg/d) | 0.15 | -0.02, 0.31 | 0.091 | 0.07 | -0.22, 0.34 | 0.664 |
| Non-heme iron (mg/d) | -0.26 | -0.41, -0.09 | 0.003** | 0.20 | -0.08, 0.46 | 0.189 |
| Zinc (mg/d) | -0.02 | -0.19, 0.15 | 0.818 | 0.21 | -0.07, 0.47 | 0.168 |
| Retinol (μg/d) | 0.01 | -0.16, 0.18 | 0.876 | 0.08 | -0.2, 0.36 | 0.587 |
| Beta carotene (μg/d) | 0.11 | -0.07, 0.27 | 0.227 | 0.27 | -0.02, 0.51 | 0.081 |
| Vitamin B1 (mg/d) | 0.04 | -0.13, 0.21 | 0.619 | 0.13 | -0.16, 0.39 | 0.413 |
| Vitamin B2 (mg/d) | -0.07 | -0.23, 0.11 | 0.459 | 0.02 | -0.27, 0.3 | 0.916 |
| Vitamin B6 (md/d) | 0.13 | -0.04, 0.29 | 0.145 | 0.00 | -0.28, 0.28 | 0.991 |

| | | | | | | |
|---|---|---|---|---|---|---|
| Vitamin B12 (µg/d) | 0.01 | -0.17, 0.18 | 0.947 | 0.03 | -0.26, 0.31 | 0.863 |
| Vitamin D (µg/d) | -0.09 | -0.26, 0.08 | 0.314 | 0.19 | -0.09, 0.45 | 0.205 |
| Vitamin E (µg/d) | -0.06 | -0.23, 0.11 | 0.482 | 0.10 | -0.18, 0.37 | 0.505 |
| Vitamin C (mg/d) | -0.15 | -0.31, 0.02 | 0.086 | -0.03 | -0.31, 0.25 | 0.836 |
| Folate total (µg/d) | -0.17 | -0.33, 0 | 0.053 | 0.19 | -0.09, 0.45 | 0.210 |
| Lycopene (µg/d) | 0.07 | -0.1, 0.24 | 0.435 | 0.03 | -0.25, 0.31 | 0.837 |
| Retinol equivalents (µg/d) | -0.10 | -0.26, 0.08 | 0.279 | 0.22 | -0.07, 0.47 | 0.154 |
| Folate equivalents (µg/d) | -0.10 | -0.26, 0.08 | 0.279 | 0.14 | -0.15, 0.41 | 0.364 |

*In the cross-sectional study, partial spearman's ρ correlations with INFLA-score were adjusted for age and BMI. In the intervention study, partial spearman's ρ correlations with ΔINFLA-score (follow-up – baseline) were adjusted for age, baseline BMI, baseline INFLA-score and the baseline value of the outcome. *P<0.05, **P<0.01.*

### Supplementary Table 6: Baseline and follow-up values of and intervention effects on adherence to DHD-guidelines, macronutrient intake and micronutrient intake

| | Cross-sectional study (n=134) | Intervention study (n=52) | | | | | | |
|---|---|---|---|---|---|---|---|---|
| | | Colchicine (n=23) | | Placebo (n=29) | | | | |
| | | Baseline | Follow-up | Baseline | Follow-up | Time x Group Effect[b] | | |
| | Mean ± SD | Mean ± SD | Mean ± SD | Mean ± SD | Mean ± SD | β (95% CI) | $R^2_{partial}$ | P-value |
| **DHD index** | | | | | | | | |
| Total (scale: 0-160) | 80.6 ± 16.0 | 81.8 ± 14.6 | 84.2 ± 18.2 | 78.4 ± 15.9 | 83.1 ± 12.5 | -3.8 (-12.3, 4.6) | 0.004 | 0.367 |
| Vegetables (scale: 0-10) | 7.8 (4.9, 10.0) | 8.1 (5.4, 10.0) | 6.2 (3.6, 9.0) | 6.5 (4.4, 10.0) | 7.7 (4.5, 10.0) | -[c] | - | - |
| Fruits (scale: 0-10) | 5.6 (3.0, 10.0) | 5.7 (3.7, 10.0) | 5.6 (2.7, 10.0) | 5.6 (3.4, 7.8) | 7.8 (4.0, 9.8) | -1.1 (-2.9, 0.8) | 0.007 | 0.246 |
| Whole grain products (scale: 0-10) | 5.9 (4.5, 8.2) | 5.5 (4.6, 6.6) | 5.3 (4.1, 7.2) | 5.3 (3.9, 7.8) | 6.2 (4.7, 8.5) | -0.1 (-1.2, 1.0) | <0.001 | 0.862 |
| Legumes (scale: 0-10) | 10.0 (2.5, 10.0) | 10.0 (2.5, 10.0) | 8.6 (2.5, 10.0) | 10.0 (7.4, 10.0) | 10.0 (5.5, 10.0) | -[c] | - | - |
| Nuts (scale: 0-10) | 3.0 (0.8, 7.3) | 2.3 (0.6, 8.0) | 2.1 (0.0, 5.8) | 1.6 (0.3, 5.0) | 3.4 (1.2, 9.7) | -[c] | - | - |
| Dairy (scale: 0-10) | 4.0 (2.0, 6.4) | 4.4 (2.5, 6.2) | 3.3 (2.0, 7.6) | 5.7 (2.0, 6.8) | 4.4 (1.4, 9.0) | -[c] | - | - |
| Fish (scale: 0-10) | 6.2 (2.9, 8.3) | 6.9 (5.0, 9.0) | 6.9 (6.2, 8.6) | 6.3 (2.9, 8.2) | 6.7 (3.3, 8.2) | -0.1 (-1.8, 1.5) | <0.001 | 0.857 |
| Tea (scale: 0-10) | 3.0 (0.5, 10.0) | 2.9 (0.3, 10.0) | 3.3 (0.5, 10.0) | 3.1 (0.3, 6.1) | 3.5 (0.7, 7.6) | -[c] | - | - |
| Fats and oils (scale: 0-10) | 6.4 (0.5, 10.0) | 10.0 (3.8, 10.0) | 10.0 (3.3, 10.0) | 5.7 (0.3, 10.0) | 7.9 (1.2, 10.0) | -[c] | - | - |
| Coffee (scale: 0-10) | 5.0 (5.0, 10.0) | 5.0 (5.0, 10.0) | 5.0 (5.0, 10.0) | 5.0 (5.0, 10.0) | 5.0 (5.0, 10.0) | -[c] | - | - |
| Red meat (scale: 0-10) | 10.0 (10.0, 10.0) | 10.0 (10.0, 10.0) | 10.0 (10.0, 10.0) | 10.0 (10.0, 10.0) | 10.0 (10.0, 10.0) | -[c] | - | - |
| Processed meat (scale: 0-10) | 1.9 (0.0, 6.6) | 3.5 (0.2, 6.6) | 4.1 (0.0, 6.7) | 1.1 (0.0, 6.0) | 0.2 (0.0, 3.6) | -[c] | - | - |
| Sugar-sweetened beverages (scale: 0-10) | 8.7 (5.9, 9.7) | 8.0 (3.7, 8.7) | 8.4 (7.8, 8.9) | 8.9 (4.8, 9.7) | 8.3 (4.4, 9.4) | -[c] | - | - |
| Alcohol (scale: 0-10) | 10.0 (10.0, 10.0) | 10.0 (10.0, 10.0) | 10.0 (10.0, 10.0) | 10.0 (10.0, 10.0) | 10.0 (10.0, 10.0) | -[c] | - | - |
| Salt (scale: 0-10) | 7.5 (5.4, 8.5) | 7.5 (5.5, 8.5) | 8.5 (7.0, 9.0) | 7.5 (4.9, 8.5) | 7.7 (5.7, 8.7) | 0.5 (-0.8, 1.8) | 0.005 | 0.431 |
| Unhealthy choices (scale: 0-10) | 0.0 (0.0, 0.7) | 0.0 (0.0, 0.8) | 0.0 (0.0, 4.7) | 0.0 (0.0, 0.0) | 0.0 (0.0, 0.3) | -[c] | - | - |
| ***Nutrients*** | | | | | | | | |
| Total energy intake (kcal/d) | 1955 ± 613 | 2069 ± 543 | 1680 ± 437 | 1924 ± 652 | 1885 ± 498 | -341 (-620, -62) | 0.023 | 0.018* |
| Protein total (g/d) | 76.5 ± 23.2 | 78.2 ± 15.9 | 67.4 ± 17.3 | 77.7 ± 22.6 | 77.5 ± 16.8 | -11 (-20, -1.7) | 0.032 | 0.021* |
| Plant protein (g/d) | 30.1 ± 10.8 | 31.0 ± 8.2 | 25.8 ± 9.4 | 29.3 ± 11.3 | 27.8 ± 8.2 | -3.3 (-7.7, 1.0) | 0.015 | 0.133 |
| Animal protein (g/d) | 46.4 ± 19.4 | 47.3 ± 14.5 | 41.7 ± 14.6 | 48.5 ± 16.3 | 49.8 ± 12.3 | -7.4 (-14, -0.8) | 0.017 | 0.030* |
| Fat total (g/d) | 85.8 ± 32.2 | 87.7 ± 27.7 | 68.5 ± 22.4 | 83.6 ± 31.7 | 82.0 ± 24.9 | -17 (-32, -2.9) | 0.053 | 0.019* |
| Saturated fat (gd) | 30.9 ± 12.9 | 32.9 ± 11.9 | 25.2 ± 9.5 | 30.4 ± 11.1 | 29.8 ± 9.3 | -7.2 (-13, -1.6) | 0.051 | 0.013* |
| Mono-unsaturated fat (g/d) | 31.5 ± 12.1 | 31.9 ± 9.7 | 24.8 ± 8.0 | 30.6 ± 12.8 | 30.2 ± 9.9 | -6.4 (-12, -1.1) | 0.049 | 0.019* |
| Poly-unsaturated(g/d) | 15.9 ± 6.8 | 15.1 ± 4.8 | 12.4 ± 4.3 | 15.3 ± 7.2 | 14.6 ± 4.9 | -2.0 (-5.0, 1.0) | 0.015 | 0.194 |
| Trans fat (g/d) | 1.5 ± 0.7 | 1.6 ± 0.7 | 1.2 ± 0.5 | 1.4 ± 0.5 | 1.5 ± 0.7 | -0.5 (-0.9, -0.1) | 0.053 | 0.020* |

| | | | | | | | |
|---|---|---|---|---|---|---|---|
| Linoleic acid (g/d) | 12.9 ± 5.7 | 12.2 ± 4.0 | 10.0 ± 3.4 | 12.5 ± 6.3 | 11.8 ± 4.0 | -1.4 (-3.9, 1.1) | 0.010 | 0.274 |
| ALA (g/d) | 1.7 ± 0.7 | 1.6 ± 0.4 | 1.3 ± 0.4 | 1.6 ± 0.8 | 1.6 ± 0.6 | -0.3 (-0.6, 0.1) | 0.022 | 0.095 |
| EPA (mg/d) | 0.1 (0.1, 0.1) | 0.1 (0.1, 0.1) | 0.1 (0.1, 0.1) | 0.1 (0.1, 0.1) | 0.1 (0.1, 0.1) | -[c] | - | - |
| DHA (g/d) | 0.1 (0.1, 0.2) | 0.1 (0.1, 0.2) | 0.1 (0.1, 0.2) | 0.1 (0.1, 0.2) | 0.1 (0.1, 0.2) | -[c] | - | - |
| Cholesterol (g/d) | 253 (188, 328) | 293 (237, 310) | 232 (182, 274) | 284 (198, 353) | 263 (199, 349) | -35 (-84, 13) | 0.007 | 0.147 |
| Carbohydrates total (g/d) | 198.7 ± 69.6 | 219.5 ± 72.5 | 177.9 ± 61.8 | 196.4 ± 75.8 | 191.5 ± 62.3 | -36 (-68, -4) | 0.038 | 0.027* |
| Sugars (g/d) | 88.2 ± 36.8 | 100.3 ± 43.5 | 81.9 ± 34.1 | 87.9 ± 38.3 | 87.9 ± 33.7 | -0.18 (-0.36, 0.01)[d] | 0.016 | 0.058 |
| Polysaccharides (g/d) | 110.4 ± 41.8 | 119.2 ± 39.8 | 96.1 ± 36.3 | 108.5 ± 47.5 | 103.5 ± 41.1 | -17 (-36, 1) | 0.027 | 0.069 |
| Fiber (g/d) | 21.5 ± 7.4 | 21.7 ± 5.2 | 18.7 ± 5.8 | 21.2 ± 8.8 | 20.7 ± 5.8 | -2.2 (-5.4, 0.9) | 0.010 | 0.157 |
| Water (ml/d) | 2333 ± 665 | 2524 ± 553 | 2196 ± 493 | 2383 ± 775 | 2325 ± 586 | -[c] | - | - |
| Alcohol (g/d) | 1.7 (0.2, 5.2) | | | | | -[c] | - | - |
| Calcium (mg/d) | 916.5 ± 329.6 | 977.2 ± 254.6 | 822.3 ± 225.6 | 922.0 ± 322.0 | 873.5 ± 296.4 | -103 (-252, 47) | 0.008 | 0.178 |
| Iron total (mg/d) | 11.0 ± 3.3 | 11.1 ± 2.1 | 9.3 ± 2.2 | 10.9 ± 3.6 | 10.6 ± 2.4 | -1.5 (-2.9, -0.1) | 0.029 | 0.042* |
| Heme iron (mg/d) | 1.0 (0.6, 1.4) | 1.1 (0.8, 1.4) | 0.8 (0.5, 1.3) | 1.0 (0.8, 1.2) | 1.2 (0.9, 1.4) | -0.3 (-0.5, -0.1) | 0.034 | 0.002** |
| Non-heme iron (mg/d) | 9.9 ± 2.9 | 10.0 ± 2.0 | 8.4 ± 2.1 | 9.9 ± 3.5 | 9.4 ± 2.4 | -1.1 (-2.5, 0.2) | 0.018 | 0.098 |
| Zinc (mg/d) | 10.0 ± 3.3 | 10.3 ± 2.2 | 8.6 ± 2.2 | 9.8 ± 2.8 | 9.9 ± 2.1 | -1.8 (-2.9, -0.7) | 0.052 | 0.002** |
| Retinol (µg/d) | 437 (290, 617) | 440 (376, 643) | 393 (317, 538) | 478 (304, 631) | 436 (308, 646) | -0.2 (-0.4, 0.1)[d] | 0.005 | 0.185 |
| Beta carotene (µg/d) | 3219 (2248, 4741) | 3656 (2530, 4982) | 1996 (1429, 4370) | 3322 (1954, 4197) | 3480 (2115, 4961) | -1183 (-2568, 203) | 0.022 | 0.093 |
| Vitamin B1 (mg/d) | 0.9 ± 0.3 | 0.9 ± 0.3 | 0.8 ± 0.2 | 1.0 ± 0.4 | 1.0 ± 0.2 | -0.2 (-0.3, 0.01) | 0.027 | 0.067 |
| Vitamin B2 (mg/d) | 1.3 ± 0.4 | 1.3 ± 0.3 | 1.1 ± 0.3 | 1.3 ± 0.4 | 1.3 ± 0.4 | -0.2 (-0.4, 0.02) | 0.016 | 0.071 |
| Vitamin B6 (md/d) | 1.6 ± 0.5 | 1.7 ± 0.4 | 1.4 ± 0.4 | 1.7 ± 0.5 | 1.6 ± 0.5 | -0.2 (-0.5, 0.1) | 0.019 | 0.102 |
| Vitamin B12 (µg/d) | 4.0 (3.2, 5.1) | 3.8 (3.2, 5.0) | 3.5 (2.9, 5.0) | 4.2 (3.5, 4.9) | 4.2 (3.7, 5.0) | - | - | - |
| Vitamin D (µg/d) | 3.0 (2.4, 4.1) | 2.9 (2.7, 3.9) | 3.2 (2.2, 3.5) | 2.7 (2.2, 4.2) | 2.9 (2.3, 4.0) | -0.3 (-0.9, 0.4) | 0.002 | 0.430 |
| Vitamin E (µg/d) | 13.5 (10.1, 17.1) | 13.5 (11.7, 15.8) | 10.4 (9.5, 13.1) | 13.1 (11.2, 18.6) | 13.6 (10.4, 16.8) | -0.21 (-0.48, 0.05)[d] | 0.024 | 0.112 |
| Vitamin C (mg/d) | 82.6 ± 33.6 | 89.5 ± 30.6 | 74.1 ± 25.9 | 91.9 ± 63.4 | 87.5 ± 36.7 | -[c] | - | - |
| Folate total (µg/d) | 232.0 ± 76.4 | 244.1 ± 57.6 | 205.1 ± 61.8 | 237.1 ± 91.6 | 216 ± 63 | -14 (-57, 30) | 0.003 | 0.532 |
| Lycopene (µg/d) | 2054 (1186, 3047) | 2082 (1294, 3021) | 1401 (1185, 2242) | 1604 (850, 3776) | 1635 (1247, 2247) | -[c] | - | - |
| Retinol equivalents (µg/d) | 780 (541, 980) | 853 (670, 1110) | 673 (542, 826) | 779 (577, 1076) | 766 (692, 915) | -0.27 (-0.52, -0.02)[d] | 0.023 | 0.037* |
| Folate equivalents (µg/d) | 253 ± 91 | 265.4 ± 69.7 | 220.7 ± 64.0 | 251 ± 92 | 232 ± 70 | -23 (-67, 21) | 0.006 | 0.302 |

*[a]Median (25th and 75th percentile) was reported due to skewed distribution of the variable.*

*[b]Intervention effects were estimated by mixed linear regression modelling with Time (follow-up, baseline) and Group (colchicine, placebo) and their interaction included as*

*fixed factors of interest, age, baseline BMI and baseline INFLA-score as covariates, and a random intercept for participant and a random slope for Time. Participants with Cook's distance >1 were removed from the analysis.* [c]*Mixed model results were not reported because the assumption of the model regarding normal distribution of the residuals was not met.* [d]*Outcome was log-transformed to correct for skewed distribution of the residuals.*

### Supplementary Table 7: Correlations between (change in) inflammation and mood, reward sensitivity, and general health outcomes

| | *Cross-sectional study (n=145), INFLA* | | | *Intervention study (n=57), ΔINFLA* | | |
|---|---|---|---|---|---|---|
| | Spearman's ρ | 95% CI | P-value | Spearman's ρ | 95% CI | P-value |
| ***Profile of mood states*** | | | | | | |
| Depressive mood[a] | -0.09 | -0.25, -0.23 | 0.305 | 0.01 | -0.26, 0.27 | 0.966 |
| Fatigue[a] | -0.07 | -0.23, -0.28 | 0.418 | 0.16 | -0.11, 0.41 | 0.269 |
| ***Hospital Anxiety and Depression Scale*** | | | | | | |
| Depression[a] | -0.08 | -0.24, -0.27 | 0.366 | 0.13 | -0.15, 0.38 | 0.386 |
| Anxiety[a] | -0.16 | -0.32, -0.33 | 0.061 | 0.09 | -0.18, 0.35 | 0.518 |
| ***Checklist Individual Strength*** | | | | | | |
| Total | -0.02 | -0.19, -0.32 | 0.771 | 0.16 | -0.12, 0.41 | 0.280 |
| Fatigue severity | 0.02 | -0.15, -0.23 | 0.802 | 0.15 | -0.12, 0.4 | 0.305 |
| Reduced activity | -0.07 | -0.23, -0.13 | 0.443 | -0.07 | -0.33, 0.2 | 0.626 |
| Reduced motivation[a] | 0.04 | -0.13, -0.19 | 0.660 | 0.20 | -0.07, 0.45 | 0.154 |
| Concentration problems[a] | -0.05 | -0.21, -0.15 | 0.579 | 0.03 | -0.24, 0.3 | 0.822 |
| ***Starkstein Apathy Scale*** | | | | | | |
| Total | 0.02 | -0.15, -0.13 | 0.826 | 0.04 | -0.23, 0.3 | 0.792 |
| ***Power of Food Scale*** | | | | | | |
| Total | 0.06 | -0.11, -0.25 | 0.512 | 0.20 | -0.07, 0.44 | 0.163 |
| Food present | 0.11 | -0.06, -0.1 | 0.200 | 0.13 | -0.14, 0.39 | 0.357 |
| Food tasted | 0.07 | -0.1, -0.11 | 0.441 | 0.28 | 0.01, 0.51 | 0.050 |
| Food available | -0.01 | -0.18, -0.06 | 0.864 | 0.12 | -0.15, 0.37 | 0.412 |
| *TEPS* | | | | | | |
| Total | -0.06 | -0.23, 0 | 0.467 | -0.06 | -0.32, 0.22 | 0.697 |
| Anticipatory reward | 0.08 | -0.09, -0.28 | 0.338 | 0.06 | -0.21, 0.33 | 0.668 |
| Consummatory reward | -0.12 | -0.28, -0.23 | 0.171 | -0.17 | -0.42, 0.1 | 0.239 |
| ***Binge Eating Scale*** | | | | | | |
| Total | -0.14 | -0.3, -0.14 | 0.093 | 0.04 | -0.23, 0.3 | 0.785 |
| *BIS/BAS* | | | | | | |
| BAS Total | -0.19 | -0.35, -0.34 | 0.025* | -0.08 | -0.34, 0.19 | 0.563 |
| BAS Reward | -0.14 | -0.3, -0.25 | 0.111 | 0.00 | -0.27, 0.27 | 0.984 |
| BAS Drive | -0.18 | -0.34, -0.25 | 0.036* | 0.01 | -0.26, 0.28 | 0.956 |
| BAS Fun | -0.09 | -0.25, -0.3 | 0.303 | -0.05 | -0.32, 0.22 | 0.709 |
| ***Kirby Monetary Choice*** | | | | | | |
| Overall k[a] (x100) | 0.06 | -0.11, -0.05 | 0.515 | 0.11 | -0.16, 0.37 | 0.430 |
| Small k[a] | 0.05 | -0.12, -0.19 | 0.582 | 0.12 | -0.16, 0.37 | 0.422 |
| Medium k[a] | 0.08 | -0.09, -0.26 | 0.353 | 0.01 | -0.26, 0.28 | 0.929 |
| Large k[a] | 0.06 | -0.11, -0.24 | 0.481 | 0.26 | -0.01, 0.49 | 0.071 |
| ***SF-36*** | | | | | | |
| Physical functioning[a] | 0.05 | -0.12, -0.15 | 0.595 | 0.10 | -0.17, 0.36 | 0.500 |
| Role – physical[a] | 0.05 | -0.12, -0.15 | 0.595 | -0.16 | -0.41, 0.11 | 0.271 |
| Pain[a] | -0.04 | -0.21, -0.12 | 0.620 | 0.01 | -0.26, 0.28 | 0.926 |
| General health | 0.04 | -0.13, -0.15 | 0.639 | -0.06 | -0.32, 0.22 | 0.703 |
| Energy/vitality | -0.06 | -0.23, -0.09 | 0.456 | -0.03 | -0.3, 0.24 | 0.832 |
| Social functioning[a] | 0.03 | -0.14, 0.20 | 0.714 | 0.09 | -0.18, 0.35 | 0.547 |
| Role – mental[a] | 0.01 | -0.15, 0.19 | 0.826 | -0.09 | -0.35, 0.18 | 0.536 |

| | | | | | | |
|---|---|---|---|---|---|---|
| Mental health | -0.03 | -0.20, 0.14 | 0.703 | -0.01 | -0.27, 0.26 | 0.959 |
| Health change | 0.02 | -0.15, 0.19 | 0.806 | 0.01 | -0.26, 0.28 | 0.927 |
| ***Pittsburgh Sleep Index*** | | | | | | |
| Global score | 0.00 | -0.17, -0.17 | 0.971 | -0.02 | -0.29, 0.25 | 0.877 |
| ***Baratt Impulsivity Scale***[b] | | | | | | |
| Total | 0.08 | -0.09, -0.35 | 0.363 | n.a. | n.a. | n.a. |
| Attention | 0.03 | -0.14, -0.12 | 0.753 | n.a. | n.a. | n.a. |
| Cognitive instability | 0.04 | -0.13, -0.11 | 0.624 | n.a. | n.a. | n.a. |
| Motor | 0.06 | -0.11, -0.13 | 0.476 | n.a. | n.a. | n.a. |
| Perseverance | -0.05 | -0.21, -0.19 | 0.592 | n.a. | n.a. | n.a. |
| Self-control | -0.03 | -0.19, -0.09 | 0.769 | n.a. | n.a. | n.a. |
| Cognitive complexity | 0.09 | -0.08, -0.13 | 0.278 | n.a. | n.a. | n.a. |
| Attentional impulsiveness | 0.05 | -0.12, -0.08 | 0.557 | n.a. | n.a. | n.a. |
| Motor impulsiveness | 0.03 | -0.13, -0.12 | 0.693 | n.a. | n.a. | n.a. |
| Non-planning impulsiveness | 0.05 | -0.12, -0.21 | 0.586 | n.a. | n.a. | n.a. |

*In the cross-sectional study, partial spearman's ρ correlations with INFLA-score were adjusted for age and BMI. In the intervention study, partial spearman's ρ correlations with ΔINFLA-score (follow-up – baseline) were adjusted for age, baseline BMI, baseline INFLA-score and the baseline value of the outcome. [a]Median (25$^{th}$, 75$^{th}$ percentile) was reported due to skewed distribution of the variable. [b]Only measured at baseline. *P<0.05.*

### Supplementary Table 8: Baseline and follow-up values of and intervention effects on mood, reward sensitivity, and general health outcomes

| | **Cross-sectional study (n=145)** | **Intervention study (n=57)** | | | | | | |
|---|---|---|---|---|---|---|---|---|
| | | **Colchicine (n=26)** | | **Placebo (n=31)** | | | | |
| | | **Baseline** | **Follow-up** | **Baseline** | **Follow-up** | **Time x Group Effect[b]** | | |
| | **Mean ± SD** | **Mean ± SD** | **Mean ± SD** | **Mean ± SD** | **Mean ± SD** | **β (95% CI)** | **$R^2_{partial}$** | **P-value** |
| ***Profile of mood states*** | | | | | | | | |
| Depressive mood[a] | 2.5 (0.0, 8.0) | 3.0 (0.0, 8.0) | 1.0 (0.0, 4.0) | 1.0 (0.0, 8.0) | 0.0 (0.0, 3.0) | -0.66 (-2.63, 1.3) | 0.002 | 0.501 |
| Fatigue[a] | 5.5 (2.0, 10.0) | 6.5 (2.0, 11.0) | 3.0 (1.0, 8.0) | 5.0 (2.0, 7.0) | 3.0 (1.0, 7.0) | -[c] | - | - |
| ***Hospital Anxiety and Depression Scale*** | | | | | | | | |
| Depression[a] | 3.0 (1.0, 5.0) | 6.0 (3.0, 9.0) | 3.0 (1.0, 4.0) | 5.0 (2.0, 7.0) | 4.0 (1.0, 5.0) | -[c] | - | - |
| Anxiety[a] | 5.5 (3.0, 8.0) | 2.5 (1.0, 5.0) | 5.5 (3.0, 8.0) | 2.0 (1.0, 6.0) | 3.0 (2.0, 7.0) | -[c] | - | - |
| ***Checklist Individual Strength*** | | | | | | | | |
| Total | 72 ± 22 | 74 ± 18 | 70 ± 21 | 66 ± 22 | 63 ± 21 | -2.15 (-10.91, 6.62) | 0.001 | 0.625 |
| Fatigue severity | 35 ± 11 | 36 ± 8 | 33 ± 12 | 33 ± 12 | 31 ± 11 | -2.22 (-7.28, 2.84) | 0.003 | 0.383 |
| Reduced activity | 6.7 ± 3.5 | 7.3 ± 3.8 | 7.1 ± 3.5 | 5.9 ± 3.3 | 5.4 ± 3.1 | 0.34 (-0.93, 1.61) | 0.001 | 0.592 |
| Reduced motivation[a] | 11.0 (7.5, 14.0) | 12.0 (9.0, 15.0) | 10.0 (8.0, 13.0) | 9.0 (6.0, 13.0) | 9.0 (6.0, 12.0) | -0.65 (-2.78, 1.48) | 0.001 | 0.541 |
| Concentration problems[a] | 17 (12, 22) | 15 (12, 23) | 15.5 (11.0, 19.0) | 15 (9, 19) | 14.0 (8.0, 20.0) | 0.28 (-2.83, 3.39) | 0.000 | 0.857 |
| ***Starkstein Apathy Scale*** | | | | | | | | |
| Total | 12.2 ± 5.1 | 13.1 ± 5.0 | 13.2 ± 4.7 | 11.0 ± 4.7 | 10.7 ± 5.2 | 0.22 (-1.41, 1.85) | 0.000 | 0.785 |
| ***Power of Food Scale*** | | | | | | | | |
| Total | 42 ± 11 | 43 ± 12 | 43 ± 12 | 45 ± 13 | 43 ± 13 | 2.23 (-2.04, 6.49) | 0.002 | 0.299 |
| Food present | 12.9 ± 3.8 | 13.6 ± 3.8 | 13.8 ± 3.9 | 14.1 ± 4.5 | 13.6 ± 4.7 | 0.7 (-1.08, 2.47) | 0.002 | 0.436 |
| Food tasted | 13.5 ± 3.7 | 13.2 ± 4.0 | 14.1 ± 3.9 | 14.5 ± 4.5 | 14.3 ± 4.6 | 0.99 (-0.85, 2.83) | 0.003 | 0.286 |
| Food available | 15.1 ± 5.4 | 16.4 ± 5.9 | 15.5 ± 5.2 | 16.3 ± 5.7 | 15.2 ± 6.3 | 0.54 (-1.5, 2.59) | 0.001 | 0.596 |
| ***TEPS*** | | | | | | | | |
| Total | 81 ± 9 | 80 ± 8 | 81 ± 8 | 83 ± 8 | 83 ± 7 | 0.53 (-3.32, 4.37) | 0.000 | 0.784 |
| Anticipatory reward | 41.4 ± 6.0 | 40.8 ± 5.5 | 41.1 ± 6.0 | 43.6 ± 5.4 | 42.8 ± 5.2 | -0.36 (-3.24, 2.52) | 0.000 | 0.803 |
| Consummatory reward | 39.3 ± 5.1 | 38.6 ± 4.3 | 40.1 ± 4.1 | 39.2 ± 4.2 | 39.9 ± 3.9 | 0.82 (-0.99, 2.63) | 0.002 | 0.368 |
| ***Binge Eating Scale*** | | | | | | | | |
| Total | 11 ± 7 | 13 ± 7 | 12 ± 6 | 11 ± 6 | 10 ± 7 | 0.14 (-1.99, 2.27) | 0.000 | 0.895 |
| ***BIS/BAS*** | | | | | | | | |
| BAS Total | 39.7 ± 4.7 | 39.3 ± 5.4 | 38.9 ± 5.1 | 40.3 ± 5.1 | 40.1 ± 5.3 | -0.07 (-1.86, 1.73) | 0.000 | 0.942 |
| BAS Reward | 15.9 ± 2.5 | 15.4 ± 2.7 | 15.5 ± 2.4 | 15.8 ± 2.5 | 15.8 ± 2.3 | 0.29 (-0.79, 1.37) | 0.001 | 0.595 |
| BAS Drive | 12.3 ± 2.1 | 12.2 ± 2.1 | 11.6 ± 2.2 | 12.5 ± 2.3 | 12.2 ± 2.2 | -0.36 (-1.16, 0.44) | 0.002 | 0.366 |
| BAS Fun | 11.5 ± 2.0 | 11.7 ± 1.9 | 11.9 ± 2.0 | 12.0 ± 2.0 | 12.1 ± 1.9 | 0.01 (-0.68, 0.7) | 0.000 | 0.981 |
| BIS | 21.6 ± 4.0 | 21.9 ± 3.6 | 21.4 ± 4.2 | 21.2 ± 3.9 | 20.8 ± 4.0 | 0.1 (-1.31, 1.52) | 0.000 | 0.883 |
| ***Kirby Monetary Choice*** | | | | | | | | |
| Overall k[a] (x100) | 0.007 (0.002, 0.015) | 0.008 (0.002, 0.020) | 0.009 (0.002, 0.020) | 0.006 (0.002, 0.015) | 0.004 (0.002, 0.015) | -[c] | - | - |
| Small k[a] | 0.01 (0.00, 0.03) | 0.010 (0.004, 0.026) | 0.010 (0.004, 0.026) | 0.010 (0.002, 0.026) | 0.004 (0.004, 0.026) | -[c] | - | - |
| Medium k[a] | 0.004 (0.002, 0.010) | 0.004 (0.002, 0.010) | 0.010 (0.002, 0.025) | 0.004 (0.002, 0.010) | 0.004 (0.002, 0.025) | -[c] | - | - |
| Large k[a] | 0.004 (0.002, 0.010) | 0.004 (0.002, 0.010) | 0.004 (0.002, 0.010) | 0.002 (0.002, 0.004) | 0.004 (0.002, 0.010) | -[c] | - | - |
| ***SF-36*** | | | | | | | | |
| Physical functioning[a] | 3. (2.0, 5.0) | 3.50 (2.00, 5.00) | 3.50 (2.00, 5.00) | 4.00 (2.00, 6.00) | 3.00 (2.00, 5.00) | -0.36 (-1.36, 0.64) | 0.002 | 0.477 |
| Role – physical[a] | 0.0 (0.0, 2.0) | 0.00 (0.00, 3.00) | 0.00 (0.00, 2.00) | 0.00 (0.00, 1.00) | 0.00 (0.00, 1.00) | -[c] | - | - |
| Pain[a] | 11 (5, 16) | 11 (5, 16) | 11 (5, 16) | 11 (5, 16) | 10 (0, 16) | 1.48 (-2.76, 5.72) | 0.002 | 0.486 |
| General health | 0.63±2.26 | 0.15 ± 2.26 | 0.96 ± 2.09 | 0.16 ± 2.45 | 1.13 ± 2.50 | 0.2 (-1.13, 1.53) | 0.000 | 0.763 |
| Energy/vitality | 10.00±1.73 | 9.77 ± 1.75 | 9.92 ± 2.12 | 10.13 ± 2.06 | 10.52 ± 1.48 | -0.13 (-1.18, 0.92) | 0.000 | 0.802 |

| | | | | | | | | |
|---|---|---|---|---|---|---|---|---|
| Social functioning[a] | 2.00 (0.00, 3.00) | 1.00 (1.00, 3.00) | 1.50 (0.00, 2.00) | 1.00 (0.00, 3.00) | 1.00 (0.00, 2.00) | -[c] | - | - |
| Role – mental[a] | 0.00 (0.00, 2.00) | 0.00 (0.00, 2.00) | 0.00 (0.00, 2.00) | 0.00 (0.00, 1.00) | 0.00 (0.00, 1.00) | -[c] | - | - |
| Mental health | 9.50±1.77 | 9.19 ± 1.55 | 9.19 ± 1.33 | 9.35 ± 1.74 | 9.23 ± 1.67 | 0.11 (-0.73, 0.94) | 0.000 | 0.797 |
| Health change | -0.17±0.87 | -0.27 ± 0.72 | 0.31 ± 0.79 | -0.06 ± 0.85 | 0.03 ± 0.87 | 0.47 (-0.05, 0.99) | 0.025 | 0.075 |
| ***Pittsburgh Sleep Index*** | | | | | | | | |
| Global score | 9.20±2.78 | 8.92 ± 2.75 | 8.92 ± 3.30 | 9.27 ± 2.03 | 7.90 ± 2.12 | 1.57 (0.25, 2.89) | 0.023 | 0.021* |
| ***Baratt Impulsivity Scale*** | | | | | | | | |
| Total | 122±16 | 130 ± 12 | n.a. | 121 ± 12 | n.a. | n.a. | n.a. | n.a. |
| Attention | 10.84±2.26 | 11.50 ± 2.12 | n.a. | 10.03 ± 1.66 | n.a. | n.a. | n.a. | n.a. |
| Cognitive instability | 6.67±1.73 | 6.85 ± 1.69 | n.a. | 6.19 ± 1.58 | n.a. | n.a. | n.a. | n.a. |
| Motor | 14.10±2.77 | 15.46 ± 1.53 | n.a. | 14.81 ± 2.82 | n.a. | n.a. | n.a. | n.a. |
| Perseverance | 7.02±1.42 | 7.23 ± 1.48 | n.a. | 7.13 ± 1.50 | n.a. | n.a. | n.a. | n.a. |
| Self-control | 11.68±2.29 | 12.58 ± 2.00 | n.a. | 11.68 ± 2.30 | n.a. | n.a. | n.a. | n.a. |
| Cognitive complexity | 10.92±2.37 | 11.35 ± 2.76 | n.a. | 10.87 ± 2.05 | n.a. | n.a. | n.a. | n.a. |
| Attentional impulsiveness | 17.5±3.4 | 18.35 ± 2.91 | n.a. | 16.23 ± 2.38 | n.a. | n.a. | n.a. | n.a. |
| Motor impulsiveness | 21.1±3.4 | 22.69 ± 2.05 | n.a. | 21.94 ± 3.16 | n.a. | n.a. | n.a. | n.a. |
| Non-planning impulsiveness | 22.6±3.6 | 23.9 ± 3.3 | n.a. | 22.5 ± 3.4 | n.a. | n.a. | n.a. | n.a. |

*[a]Median (25th and 75th percentile) was reported due to skewed distribution of the variable. [b]Intervention effects were estimated by mixed linear regression modelling with Time (follow-up, baseline) and Group (colchicine, placebo) and their interaction included as fixed factors of interest, age, baseline BMI and baseline INFLA-score as covariates, and a random intercept for participant and a random slope for Time. [c]Mixed model results were not reported because the assumptions of the model regarding normal distribution of the residuals were not met.*

**Supplementary Table 9: Mixed linear regression models to study the association between (change in) task-related effort aversion and effort-related food intake**

| | Cross-sectional study (n=130) | | | Intervention study (n=48) | | |
|---|---|---|---|---|---|---|
| **Predictor** | **β (95% CI)** | **$R^2_{partial}$** | **p-value** | **β (95% CI)** | **$R^2_{partial}$** | **p-value** |
| Effort (high vs. low) | -0.49 (-0.62, -0.37) | 0.055 | <0.001*** | -0.53 (-0.67, -0.38) | 0.058 | <0.001*** |
| Liking (SD) | 0.44 (0.37, 0.50) | 0.152 | <0.001*** | 0.49 (0.41, 0.57) | 0.174 | <0.001*** |
| EffortAversion_t1 (Z-score) | -0.04 (-0.11, 0.04) | 0.001 | 0.332 | -0.06 (-0.20, 0.09) | 0.002 | 0.429 |
| Age (SD) | -0.03 (-0.10, 0.05) | 0.001 | 0.456 | -0.02 (-0.16, 0.11) | 0.001 | 0.706 |
| BMI_t1 (SD) | 0.00 (-0.08, 0.08) | <0.001 | 0.967 | -0.06 (-0.19, 0.08) | 0.003 | 0.412 |
| INFLA_t1 (SD) | - | - | - | -0.09 (-0.33, 0.16) | 0.002 | 0.477 |
| ΔEffortAversion (b) | - | - | - | 0.02 (-0.12, 0.16) | <0.001 | 0.742 |
| Time (t1, t2) | - | - | - | 0.14 (-0.01, 0.29) | 0.004 | 0.062 |
| Effort x Liking | -0.03 (-0.15, 0.010) | <0.001 | 0.680 | -0.18 (-0.33, -0.03) | 0.007 | 0.017* |
| EffortAversion_t1 x Effort | 0.03 (-0.10, 0.05) | 0.001 | 0.639 | - | - | - |
| EffortAversion_t1 x Liking | 0.09 (0.03, 0.16) | 0.008 | 0.004** | - | - | - |
| EffortAversion_t1 x Effort x Liking | -0.15 (-0.28, -0.03) | 0.005 | 0.018* | - | - | - |
| ΔEffortAversion x Time | - | - | - | 0.01 (-0.13, 0.16) | <0.001 | 0.872 |
| Time x Liking | - | - | - | 0.10 (-0.05, 0.24) | 0.002 | 0.191 |
| Effort x Time | - | - | - | -0.17 (-0.46, 0.12) | 0.002 | 0.250 |
| ΔEffortAversion x Effort | - | - | - | -0.04 (-0.18, 0.10) | <0.001 | 0.584 |
| ΔEffortAversion x Liking | - | - | - | -0.09 (-0.16, -0.02) | 0.008 | 0.012* |
| Effort x Time x Liking | - | - | - | -0.11 (-0.18, 0.40) | 0.001 | 0.471 |
| ΔEffortAversion x Effort x Liking | - | - | - | 0.05 (-0.09, 0.19) | 0.001 | 0.450 |
| ΔEffortAversion x Effort x Time | - | - | - | -0.14 (-0.43, 0.15) | 0.001 | 0.338 |
| ΔEffortAversion x Liking x Time | - | - | - | -0.04 (-0.18, 0.09) | <0.001 | 0.550 |
| ΔEffortAversion x Effort x Liking x Time | - | - | - | 0.10 (-0.17, 0.37) | 0.001 | 0.464 |

*Results are obtained using mixed linear regression analysis. The outcome Caloric Intake was log-transformed to correct of skewed distribution of the residuals. All continuous variables in the model were standardized. CI, confidence interval. *P<0.05, **P<0.01 ***P<0.001*

### Supplementary Table 10: Mixed linear regression models to study the association between (change in) effort-related dmPFC signal and effort-related food intake

| | Cross-sectional study (n=130) | | | Intervention study (n=48) | | |
|---|---|---|---|---|---|---|
| **Predictor** | **β (95% CI)** | **$R^2_{partial}$** | **p-value** | **β (95% CI)** | **$R^2_{partial}$** | **p-value** |
| Effort (high vs. low) | -0.50 (-0.62, -0.38) | 0.055 | <0.001*** | -0.53 (-0.68, -0.39) | **0.059** | **<0.001*** ** |
| Liking (SD) | 0.44 (0.38, 0.50) | 0.151 | <0.001*** | 0.49 (0.41, 0.56) | 0.165 | <0.001*** |
| dmPFC_signal_t1 (Z-score) | 0.05 (-0.03, 0.12) | 0.002 | 0.191 | -0.10 (-0.28, 0.08) | 0.005 | 0.253 |
| Age (SD) | -0.03 (-0.10, 0.05) | 0.001 | 0.465 | -0.07 (-0.21, 0.06) | 0.005 | 0.291 |
| BMI_t1 (SD) | 0.00 (-0.08, 0.07) | <0.001 | 0.905 | -0.06 (-0.19, 0.08) | 0.003 | 0.389 |
| INFLA_t1 (SD) | - | - | - | -0.19 (-0.44, 0.05) | 0.009 | 0.117 |
| ΔdmPFC_signal (b) | - | - | - | -0.16 (-0.35, 0.02) | 0.011 | 0.079 |
| Time (t1, t2) | - | - | - | 0.15 (0.00, 0.29) | 0.005 | 0.045* |
| Effort x Liking | -0.03 (-0.15, 0.10) | <0.001 | 0.671 | -0.18 (-0.32, -0.03) | 0.006 | 0.021* |
| dmPFC_signal_t1 x Effort | -0.08 (-0.20, 0.04) | 0.002 | 0.200 | - | - | - |
| dmPFC_signal_t1 x Liking | -0.01 (-0.08, 0.05) | <0.001 | 0.726 | - | - | - |
| dmPFC_signal_t1 x Effort x Liking | -0.01 (-0.14, 0.12) | <0.001 | 0.874 | - | - | - |
| ΔdmPFC_signal x Time | - | - | - | 0.12 (-0.02, 0.26) | 0.003 | 0.106 |
| Time x Liking | - | - | - | 0.13 (-0.01, 0.28) | 0.004 | 0.077 |
| Effort x Time | - | - | - | -0.18 (-0.46, 0.11) | 0.002 | 0.224 |
| ΔdmPFC_signal x Effort | - | - | - | 0.05 (-0.09, 0.19) | 0.001 | 0.506 |
| ΔdmPFC_signal x Liking | - | - | - | 0.00 (-0.08, 0.07) | <0.001 | 0.963 |
| Effort x Time x Liking | - | - | - | 0.06 (-0.23, 0.35) | <0.001 | 0.697 |
| ΔdmPFC_signal x Effort x Liking | - | - | - | -0.05 (-0.19, 0.10) | <0.001 | 0.541 |
| ΔdmPFC_signal x Effort x Time | - | - | - | 0.05 (-0.24, 0.33) | <0.001 | 0.749 |
| ΔdmPFC_signal x Liking x Time | - | - | - | 0.09 (-0.05, 0.24) | 0.002 | 0.201 |
| ΔdmPFC_signal x Effort x Liking x Time | - | - | - | 0.15 (-0.14, 0.43) | 0.001 | 0.305 |

*Results are obtained using mixed linear regression analysis. The outcome Caloric Intake was log-transformed to correct of skewed distribution of the residuals. All continuous variables in the model were standardized. CI, confidence interval. *P<0.05, **P<0.01 ***P<0.001*

**Supplementary Table 11: Associations between task-related effort aversion and motivational EMA components**

| Predictors (time t-1) | Outcome (time t) | β / OR | 95% CI | $R^2_{partial}$ | P-value |
|---|---|---|---|---|---|
| ***Associations with inflammation*** | | | | | |
| Effort aversion | Anticipation (scale: 1-7) | 0.00 | -0.06, 0.07 | <0.001 | 0.886 |
| | Activity completion[a] (yes vs. no) | 1.09 | 0.97, 1.25 | 0.002 | 0.150 |
| | Log(Subjective effort (scale: 1-7))[b] | -0.02 | -0.06, 0.01 | 0.006 | 0.184 |
| | Activity type[a] (high vs. low effort) | 0.97 | 0.92, 1.02 | 0.001 | 0.259 |

*Associations between Effort aversion and EMA outcomes are obtained using mixed linear regression analysis. [a]Odds ratios are reported for dichotomous outcome variables. [b]Subjective Effort was log-transformed to correct of skewed distribution of the residuals. CI, confidence interval; dmPFC, dorsomedial prefrontal cortex; EMA, ecological momentary assessment, OR, odds ratio. *P<0.05.*

### Supplementary Table 12: Associations between effort-related dmPFC signal and motivational EMA components

| Predictors (time t-1) | Outcome (time t) | β / OR | 95% CI | $R^2_{partial}$ | P-value |
|---|---|---|---|---|---|
| ***Associations with inflammation*** | | | | | |
| dmPFC signal | Anticipation (scale: 1-7) | 0.09 | -0.07, 0.26 | 0.003 | 0.261 |
| | Activity completion[a] (yes vs. no) | 1.01 | 0.73, 1.40 | <0.001 | 0.960 |
| | Log(Subjective effort (scale: 1-7))[b] | -0.06 | -0.13, 0.01 | 0.009 | 0.082 |
| | Activity type[a] (high vs. low effort) | 0.99 | 0.87, 1.13 | <0.001 | 0.854 |

*Associations between dmPFC signal and EMA outcomes are obtained using mixed linear regression analysis. All continuous predictors in the model were standardized. [a]Odds ratios are reported for dichotomous outcome variables. [b]Subjective Effort was log-transformed to correct of skewed distribution of the residuals. CI, confidence interval; dmPFC, dorsomedial prefrontal cortex; EMA, ecological momentary assessment, OR, odds ratio. *P<0.05.*

### Supplementary Table 13: Correlations between (change in) task-related effort aversion and diet quality

| | ***Cross-sectional study (n=121), effortAversion*** | | | ***Intervention study (n=45), ΔEffortAversion*** | | |
|---|---|---|---|---|---|---|
| | **Spearman's ρ** | **95% CI** | **P-value[a]** | **Spearman's ρ** | **95% CI** | **P-value[a]** |
| Total | 0.07 | -0.11. 0.25 | 0.446 | -0.05 | -0.34. 0.24 | 0.732 |
| Vegetables | 0.00 | -0.18. 0.18 | 0.988 | -0.35 | -0.58. -0.07 | 0.022 |
| Fruits | 0.01 | -0.17. 0.19 | 0.944 | -0.12 | -0.39. 0.18 | 0.458 |
| Whole grain products | -0.01 | -0.19. 0.17 | 0.923 | 0.06 | -0.23. 0.34 | 0.717 |
| Legumes | -0.13 | -0.3. 0.05 | 0.154 | 0.06 | -0.23. 0.34 | 0.720 |
| Nuts | 0.03 | -0.15. 0.21 | 0.753 | 0.06 | -0.24. 0.34 | 0.725 |
| Dairy | 0.00 | -0.18. 0.18 | 0.973 | 0.02 | -0.27. 0.31 | 0.885 |
| Fish | 0.10 | -0.08. 0.28 | 0.254 | -0.15 | -0.42. 0.15 | 0.350 |
| Tea | -0.02 | -0.19. 0.16 | 0.865 | -0.04 | -0.32. 0.25 | 0.802 |
| Fats and oils | 0.07 | -0.11. 0.25 | 0.429 | 0.08 | -0.21. 0.36 | 0.595 |
| Coffee | -0.10 | -0.27. 0.08 | 0.283 | 0.08 | -0.21. 0.36 | 0.591 |
| Red meat | 0.17 | -0.01. 0.34 | 0.068 | 0.06 | -0.23. 0.34 | 0.714 |
| Processed meat | 0.11 | -0.07. 0.28 | 0.235 | -0.13 | -0.4. 0.17 | 0.420 |
| Sugar-sweetened beverages | 0.13 | -0.05. 0.3 | 0.170 | -0.11 | -0.38. 0.19 | 0.502 |
| Alcohol | -0.15 | -0.32. 0.03 | 0.096 | 0.16 | -0.13. 0.43 | 0.298 |
| Salt | 0.17 | -0.01. 0.34 | 0.065 | -0.05 | -0.33. 0.24 | 0.765 |
| Unhealthy choices | 0.06 | -0.12. 0.24 | 0.489 | -0.03 | -0.31. 0.26 | 0.852 |

*In the cross-sectional study, partial spearman's ρ correlations with INFLA-score were adjusted for age and BMI. In the intervention study, partial spearman's ρ correlations with ΔINFLA-score (follow-up – baseline) were adjusted for age, baseline BMI, baseline INFLA-score and the baseline value of the outcome. *P<0.05, **P<0.01.*

### Supplementary Table 14: Correlations between (change in) effort-related dmPFC signal and diet quality

| | Cross-sectional study (n=121). dmPFC_signal | | | Intervention study (n=45). ΔdmPFC_signal | | |
|---|---|---|---|---|---|---|
| | Spearman's ρ | 95% CI | P-value | Spearman's ρ | 95% CI | P-value |
| Total | 0.00 | -0.17, 0.18 | 0.959 | 0.02 | -0.25, 0.29 | 0.881 |
| Vegetables | -0.02 | -0.19, 0.16 | 0.824 | 0.00 | -0.27, 0.28 | 0.984 |
| Fruits | 0.04 | -0.13, 0.22 | 0.624 | 0.16 | -0.12, 0.41 | 0.285 |
| Whole grain products | -0.16 | -0.33, 0.01 | 0.073 | 0.21 | -0.06, 0.46 | 0.144 |
| Legumes | -0.08 | -0.25, 0.1 | 0.389 | 0.05 | -0.23, 0.32 | 0.732 |
| Nuts | 0.12 | -0.05, 0.29 | 0.167 | 0.04 | -0.23, 0.31 | 0.768 |
| Dairy | -0.12 | -0.29, 0.06 | 0.193 | 0.06 | -0.21, 0.33 | 0.662 |
| Fish | -0.03 | -0.2. 0.15 | 0.740 | -0.27 | -0.51, 0.00 | 0.060 |
| Tea | 0.06 | -0.12, 0.23 | 0.541 | -0.09 | -0.36, 0.18 | 0.526 |
| Fats and oils | 0.07 | -0.10, 0.24 | 0.424 | 0.04 | -0.24, 0.31 | 0.787 |
| Coffee | 0.10 | -0.08, 0.27 | 0.272 | 0.05 | -0.22, 0.32 | 0.721 |
| Red meat | 0.12 | -0.06, 0.29 | 0.188 | -0.23 | -0.48, 0.04 | 0.109 |
| Processed meat | -0.17 | -0.34, 0.00 | 0.057 | -0.18 | -0.43, 0.1 | 0.230 |
| Sugar-sweetened beverages | 0.10 | -0.08, 0.27 | 0.260 | 0.05 | -0.22, 0.32 | 0.712 |
| Alcohol | 0.00 | -0.17, 0.18 | 0.986 | 0.05 | -0.23, 0.32 | 0.745 |
| Salt | -0.12 | -0.29, 0.06 | 0.182 | -0.35 | -0.57, -0.09 | 0.014* |
| Unhealthy choices | -0.13 | -0.30, 0.04 | 0.136 | -0.12 | -0.38, 0.16 | 0.435 |

*In the cross-sectional study, partial spearman's ρ correlations with INFLA-score were adjusted for age and BMI. In the intervention study, partial spearman's ρ correlations with ΔINFLA-score (follow-up – baseline) were adjusted for age, baseline BMI, baseline INFLA-score and the baseline value of the outcome. *P<0.05, **P<0.01.*

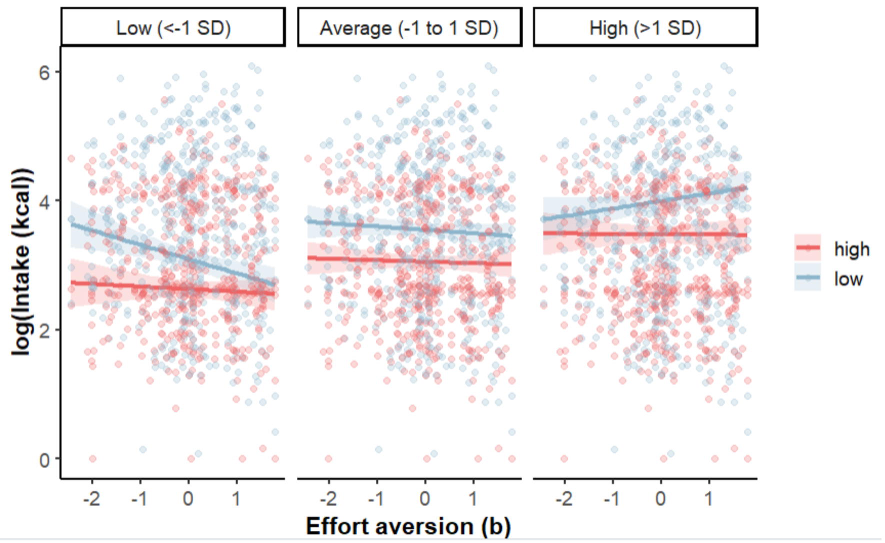


**Supplementary Fig. 1: Interaction between task-related effort aversion, effort, and liking predicting caloric food intake, as tested using linear mixed-effects modelling**
